# LAMP-coupled CRISPR-Cas12a assays for upgrading molecular detection of *Leishmania* infections

**DOI:** 10.1101/2025.08.08.25333281

**Authors:** Eva Dueñas, Ingrid Tirado, Percy Huaihua, Ariana Parra del Riego, Luis Cabrera-Sosa, Jose A. Nakamoto, María Cruz, Carlos M. Restrepo, Jorge Arévalo, Vanessa Adaui

## Abstract

**Background:** Tegumentary leishmaniasis is a parasitic disease endemic in the Americas. Its clinical management and control rely on early and accurate diagnosis and adequate treatment. PCR-based molecular diagnostics offer high sensitivity and specificity over microscopy or culture but are less accessible in low-resource settings. New molecular tools for detecting Leishmania infections are needed in rural endemic regions. A promising tool harnessing CRISPR-Cas technology enables highly specific and sensitive detection of nucleic acid targets, offering an exciting potential for portable molecular diagnostics. Previously, we developed CRISPR-Cas12a-based assays coupled to PCR preamplification for Leishmania detection. Here, we adapted our assays, which target the 18S rDNA and kinetoplast DNA (kDNA) minicircles, by replacing PCR with loop-mediated isothermal amplification (LAMP).

**Methodology/Principal Findings:** LAMP-CRISPR assays were optimized for fluorescence-based and lateral flow readouts. The assays could detect as low as 0.2 genome equivalents per reaction using *L. braziliensis* M2904 strain genomic DNA. The kDNA assay reliably detected all tested species of the *Leishmania* (*Viannia*) subgenus, while the 18S assay showed pan-*Leishmania* detection capability. There was no cross-reactivity with other protozoan (*Trypanosoma cruzi* and *Plasmodium falciparum*) and bacterial (*Mycobacterium tuberculosis*) pathogen DNA, or with human DNA. When applied to 90 clinical samples (skin lesions) from the Cusco region of Peru and compared to kDNA real-time PCR, LAMP-CRISPR assays with a fluorescence readout achieved a sensitivity of 90.9% for kDNA and 72.7% for 18S rDNA, both with 100% specificity. Overall, lateral flow strip results agreed with fluorescence-based detection in 18 tested samples, with one discrepancy observed in the 18S assay associated with low parasite load.

**Conclusions/Significance:** These new assays, being amenable to further simplification and optimization for their adoption in low-resource settings, hold promise as a new generation of accurate molecular tools for leishmaniasis diagnosis and surveillance, supporting One Health strategies for disease control.

**Author Summary:** Tegumentary leishmaniasis affects poverty-related populations in the Americas and encompasses skin and mucosal lesions that can cause disfigurement and social stigma. The disease is caused by several species of the protozoan parasite Leishmania. PCR-based molecular diagnostics are currently the most sensitive and specific diagnostic tools. Yet, these require specialized infrastructure and trained personnel that are not readily available in low-resource settings. New tools are required to meet the diagnostic needs in rural endemic areas. A promising tool leveraging CRISPR-Cas technology enables cost-effective, in vitro nucleic acid detection, paving the way for diagnostic solutions that could be made available to patients at, or near, the point of care. Here, we harnessed the CRISPR-Cas12a system combined with loop-mediated isothermal amplification (LAMP) to develop assays capable of detecting multiple Leishmania species of medical importance. Our assays employ multi-copy targets widely used in molecular diagnostics: the 18S rDNA for pan-Leishmania detection and a kDNA minicircle region conserved among L. (Viannia) species. Results can be read with either fluorescence detection or lateral flow strips. Both assays showed satisfying performance in both analytical validation and clinical sample testing under laboratory conditions. These new tools show promise to improve diagnosis and surveillance of leishmaniasis.

## Introduction

Leishmaniasis is one of the most serious neglected infectious diseases, with 0.7 to 1 million new cases each year and between 20,000 and 30,000 deaths [1, 2]. As a vector-borne disease, leishmaniasis poses a substantial threat to millions of individuals in regions where the disease is endemic, which include Asia, South and Central America, Africa and the Mediterranean basin [3]. Leishmaniasis results from an infection with protozoan parasites of the genus *Leishmania* that are transmitted to humans (and other mammals) through the bite of infected sandflies and manifests in two main clinical forms: cutaneous (or tegumentary) and visceral leishmaniasis, each presenting unique challenges in terms of diagnosis, treatment, and control [4, 5]. In the Americas, tegumentary leishmaniasis is widely distributed in 18 endemic countries and comprises a spectrum of clinical conditions ranging from localized skin ulcers to severe disfiguring mucosal lesions [5]. This clinical pleomorphism is largely dependent on the immune responses of the human host as well as on factors intrinsic to the different infecting *Leishmania* species [6]. Diverse *Leishmania* species from the *Viannia* and *Leishmania* subgenera circulate in several Central and South American countries, with the former being responsible for the majority of leishmaniasis cases [4]. While efforts directed toward the development of effective prophylactic and therapeutic vaccines against human leishmaniasis are ongoing, early and accurate diagnosis paired with timely chemotherapy remain the most vital tools in the fight against the leishmaniases [7].

The diagnostic tools currently available for the detection of *Leishmania* spp. infections include direct smear examination (light microscopy), *in vitro* culture, immunological and molecular techniques [8]. Among the nucleic acid amplification techniques (NAATs), the polymerase chain reaction (PCR) and its variants, such as real-time PCR, currently provide the most sensitive and specific diagnostic tools available, and several assays have been developed for the detection and identification of *Leishmania* species [9, 10]. While accessible in research and reference laboratories, PCR-based methods may not be readily available in rural endemic areas since they require infrastructure, resources, and technical expertise [11]. Because of this, (near) point-of-care (PoC) diagnostic tests have become a noteworthy concept, as they would allow easier, faster and less expensive diagnostic alternatives that need only minimal laboratory setup [12]. Advancements towards simplifying molecular diagnostic tools gave rise to multiple isothermal NAATs, such as loop-mediated isothermal amplification (LAMP) and recombinase polymerase amplification (RPA), which hold potential to decentralize molecular diagnostic testing [13]. Of these, LAMP has gained prominence due to its simplicity and effectiveness in detecting nucleic acid sequences with high specificity. LAMP is different from the traditional PCR as it operates at a constant temperature (60-65°C), eliminating the need for complex thermal cycling equipment [14, 15]. The LAMP reaction uses 4 to 6 primers that target specific DNA sequences, along with the Bst DNA polymerase, an enzyme that displays a strong strand displacement activity [14]. LAMP amplifies the target DNA producing a concatenated DNA product with stem-loop DNA structures, yielding up to 10^9^ copies in less than an hour [14]. The results can be easily visualized, e.g. on the basis of the turbidity caused by reaction by-products [16], or with the use of sequence-specific probes [17, 18]. Due to its user-friendly nature and rapidity, LAMP holds great promise as a tool for the diagnosis of infectious diseases in resource-limited settings [19]. Several studies have reported LAMP assays for genus-, complex-or species-specific detection of *Leishmania*; these achieved high sensitivity and specificity and showed good diagnostic performance for human cutaneous leishmaniasis (CL) and visceral leishmaniasis (VL) (reviewed in [20, 21]). A commercially available LAMP kit (Loopamp™ *Leishmania* Detection Kit, developed by Eiken Chemical, Japan) to detect *Leishmania* DNA has also been evaluated for the diagnosis of human CL and VL and showed comparable performance to PCR-based methods [22].

Clustered regularly interspaced short palindromic repeats (CRISPR)/CRISPR-associated proteins (Cas) technology has recently been harnessed as a tool for molecular detection and the development of CRISPR-based diagnostics [23], based on the discovery that once Cas12a and Cas13a enzymes bound to CRISPR RNA (crRNA) recognize specific nucleic acid targets, they become activated, thereby catalyzing cleavage of both the targets (*cis*-cleavage) as well as non-target nucleic acids (*trans*-cleavage) [24, 25]. This collateral *trans*-cleavage activity can be applied for nucleic acid detection by coupling it with a reporter molecule. For fluorescence-based assays, the reporter molecule consists of a DNA or RNA oligomer carrying a fluorophore and a quencher. So, upon target recognition, reporter cleavage by the activated Cas enzyme generates a detectable fluorescence signal that indicates that the target nucleic acid sequence is present in a sample. Alternatively, visualization of the Cas detection reaction can be achieved by a lateral flow strip readout using a reporter molecule labeled with FAM or digoxin and biotin [26, 27]. CRISPR-based assay platforms reported thus far enable a highly sensitive detection of nucleic acids in the attomolar range when combined with a target preamplification step using either isothermal NAATs (e.g. DETECTR [24] and SHERLOCK [25]) or PCR-based NAATs (e.g.

HOLMES [28]). Notwithstanding that this two-step CRISPR-based assay format increases assay complexity and poses the risk of cross-contamination during sample transfer to the CRISPR reaction, it is a convenient way to pursue in the first development phase of CRISPR-based assays to demonstrate their potential for further assay development [29]. More recent efforts aim for amplification-free detection of nucleic acid targets with CRISPR, and the proof-of-concept was demonstrated for direct detection of pathogen RNA and DNA with quantitative ability [30–33].

The combination of CRISPR-Cas and LAMP technologies has advanced the progress of nucleic acid-based PoC diagnostic test development. Coupling LAMP reactions to downstream CRISPR-based detection improves specificity [19, 23], since LAMP is prone to non-specific amplification [34]. Integrated LAMP-CRISPR assays have been developed for the detection of several pathogens, including viruses (e.g. SARS-CoV-2 [26, 35–37], human papillomavirus [38]), bacteria (e.g. *Mycobacterium tuberculosis* [39], *Klebsiella pneumoniae* [40]), and fungi (e.g. *Verticillium dahliae* [41]). CRISPR-based assays have also been developed for the detection of parasites of medical and veterinary importance, such as the protozoa *Plasmodium* spp. [29, 42], *Toxoplasma gondii* [43], *Leishmania* spp. [44–47], *Trypanosoma brucei* [48, 49], and *Trypanosoma cruzi* [50]; and the trematode *Schistosoma* species [51, 52]. These assays, aiming at sensing parasite nucleic acid sequences in biological specimens, mostly combined CRISPR with RPA preamplification. Applications of LAMP-CRISPR for parasite detection are increasing, as illustrated by the detection of *Schistosoma* infection in host samples [53], the *in silico* design of LAMP primers and Cas12a crRNA sequences for identifying genetic polymorphisms associated with drug resistance in *P. falciparum* infections [54], the detection of *T. gondii* in environmental samples [55], and the detection of *T. cruzi* in infants for diagnosing acute congenital infection [56].

At present, leishmaniasis diagnosis in endemic areas is impaired mainly by the lack of infrastructure, equipment, trained human resources, and low access (if any) to effective diagnostic tools. New molecular diagnostic tools that are accurate, easy to use, accessible and affordable for use in resource-limited settings are required. In previous work, we developed CRISPR-Cas12a-based assays coupled to PCR preamplification for detection of *Leishmania* spp. in clinical samples [45]. Here, we have adapted our assays, which target the *Leishmania* 18S ribosomal RNA gene (18S rDNA) and minicircle kinetoplast DNA (kDNA), by replacing PCR with LAMP in the preamplification step, to facilitate the road towards the development of a nucleic acid-based near-PoC diagnostic tool for leishmaniasis. We optimized LAMP-CRISPR assay conditions with a fluorescence signal readout in a plate reader as well as a visual readout on lateral flow strips. We report the evaluation of LAMP-CRISPR assay diagnostic performance against a kDNA real-time PCR assay on stored clinical samples from the Cusco region, a highly endemic area for tegumentary leishmaniasis in south-central Peru where *Leishmania* species of the *Viannia* subgenus circulate [57, 58].

## Methods

### DNA samples from reference strains of *Leishmania* and other microbial pathogens

Genomic DNA (gDNA) samples extracted from cultured promastigotes of reference strains of *Leishmania* spp. analyzed here were retrieved from the DNA biobank of the *Leishmania* research group at the Molecular Epidemiology Unit of the Instituto de Medicina Tropical Alexander von Humboldt (IMTAvH), Universidad Peruana Cayetano Heredia (UPCH) in Lima, Peru, and from the collection hosted by the Institute for Scientific Research and High Technology Services (INDICASAT-AIP) in Panama. gDNA samples isolated from cultured laboratory reference strains of other microbial pathogens (*Trypanosoma cruzi*, *Plasmodium falciparum*, *Mycobacterium tuberculosis*) were provided by the Infectious Diseases Research Laboratory at the UPCH and INDICASAT-AIP. The designation and source of the strains whose gDNA were analyzed in this study are listed in Table 1.

**Table 1.** Reference strains used in this work.

| Strain name | WHO code<sup>‡</sup> | Source |
| --- | --- | --- |
| *Leishmania (Viannia) braziliensis* | MHOM/BR/75/M2904 | IMTA<sub>v</sub>H-UPCH |
| *Leishmania (Viannia) braziliensis* | MHOM/BR/75/M2903 | INDICASAT-AIP |
| *Leishmania (Viannia) braziliensis* | MHOM/PE/91/LC2043 | IMTA<sub>v</sub>H-UPCH |
| *Leishmania (Viannia) braziliensis* | MHOM/PE/91/LC2177 | IMTA<sub>v</sub>H-UPCH |
| *Leishmania (Viannia) peruviana* | MHOM/PE/90/HB86 | IMTA<sub>v</sub>H-UPCH |
| *Leishmania (Viannia) peruviana* | MHOM/PE/90/LCA08 | IMTA<sub>v</sub>H-UPCH |
| *Leishmania (Viannia) peruviana* | MHOM/PE/2005/WR-2771 | INDICASAT-AIP |
| *Leishmania (Viannia) guyanensis* | MHOM/BR/1975/M4147 | INDICASAT-AIP |
| *Leishmania (Viannia) lainsoni* | MHOM/BR/1981/M6426 | INDICASAT-AIP |
| *Leishmania (Viannia) panamensis* | MHOM/PA/2018/BD-02 | INDICASAT-AIP |
| *Leishmania (Leishmania) amazonensis* | MHOM/BR/1973/M2269 | INDICASAT-AIP |
| *Leishmania (Leishmania) mexicana* | MHOM/BZ/1982/BEL21 | INDICASAT-AIP |
| *Leishmania (Leishmania) aristidesi* | MORY/PA/1969/GML | INDICASAT-AIP |
| *Leishmania (Leishmania) major* | MHOM/SA/1991/WR-1088 | INDICASAT-AIP |
| *Leishmania (Leishmania) infantum* | MCAN/BR/1997/P142 | INDICASAT-AIP |
| *Leishmania (Leishmania) donovani* | MHOM/IN/2006/WR-2801 | INDICASAT-AIP |
| *Trypanosoma cruzi* | Sylvio X10 | UPCH |
| *Trypanosoma cruzi* | VT-1 | UPCH |
| *Trypanosoma cruzi* | Tulahuen LacZ clone C4 | UPCH |
| *Plasmodium falciparum* | HB3 | INDICASAT-AIP |
| *Mycobacterium tuberculosis* | PA155-18C | INDICASAT-AIP |

<sup>‡</sup>The World Health Organization (WHO) designation code for *Leishmania* strains is as follows: host (M for Mammalia; HOM = *Homo sapiens*; CAN = *Canis familiaris*; ORY = *Oryzomys capito*)/country of origin (BR = Brazil; BZ = Belize; IN = India; PA = Panama; PE = Peru; SA = Saudi Arabia)/year of isolation/name of strain.

At the IMTAvH and UPCH, DNA was isolated using the High Pure PCR Template Preparation Kit (Roche, Mannheim, Germany). At the INDICASAT-AIP, the Wizard® Genomic DNA Purification Kit (Promega, Madison, WI, USA) was used. DNA was isolated according to the manufacturer’s protocol, suspended in elution buffer or TE buffer and stored at −20°C until use.

### Patient DNA samples Ethics statement

This study used anonymized stored DNA samples that were assigned a code and had been isolated from different human skin ulcerative lesion specimen types, including biopsy, lancet scraping, cytological brush, swab, and filter paper. The respective patients had provided written informed consent for future research use of their specimens and clinical data. Patients with clinically suspected CL were recruited at the Hospital Nacional Adolfo Guevara Velasco (HNAGV) in Cusco, a region in Peru with endemic circulation of *Leishmania* (*Viannia*) parasites, most frequently *L. braziliensis*, *L. guyanensis* and *L. lainsoni* [57, 58], as part of another study between the HNAGV and UPCH (contract 095-2018-FONDECYT-BM-IADT-AV to JA, financed by CONCYTEC and The World Bank). The study protocol and informed consent (registration number 103155) were reviewed and approved by the ethics committee of the UPCH (IRB approval letter 063-05-19 dated 01/30/2019, latest renewed on 05/02/2023 with letter R-149-17-23).

### DNA extraction from clinical samples prior to this study

DNA from clinical samples was isolated with the High Pure PCR Template Preparation Kit (Roche), according to the manufacturer’s protocol, suspended in elution buffer and stored at −20°C until use.

### LAMP primer design

The 18S rDNA gene (TriTrypDB ID: LbrM.27.2.208540; at https://tritrypdb.org/tritrypdb/) and kDNA minicircles (GenBank accession no. KY698803.1) from *L*. *braziliensis* MHOM/BR/75/M2904 were selected for LAMP primer design. FASTA sequences up to 200 bp downstream and upstream to previously reported crRNA target recognition sites for each gene [45] were uploaded to PrimerExplorer V5 software (Eiken Chemical Co., Ltd). Three sets of external (F3 and B3) and internal (FIP and BIP) primers were obtained for 18S rDNA, but only one for kDNA. To select the final 18S primer set, the following features were considered: the less likelihood of primer dimer formation (highest ΔG value), the 3’ end stability for F2 and B2 (ΔG ≤ −4 kcal/mol), the 5’ end stability for F1c and B1c (ΔG ≤ −4 kcal/mol) and the less overlap between primers and the recognition site. After that, loop primers (LF and LB) for both target genomic regions were also designed using the PrimerExplorer V5 software. Final primer sets were ordered from Macrogen Inc. (Seoul, South Korea) and are listed in the S1 Table.

To investigate the *in silico* specificity of the designed LAMP primers, we downloaded all publicly available DNA sequences for each target from NCBI in May 2025. For *Leishmania* kDNA minicircle sequence analysis, a total of 1,060 sequences from the *L*. (*Viannia*) subgenus and 965 sequences from the *L*. (*Leishmania*) subgenus were retained and analyzed following a quality filtering process. Sequences that were excluded contained ambiguous base pairs (e.g. ‘N’) exceeding 10% of the total length or did not cover the primer binding sites. The eight LAMP primer binding regions and the crRNA target site were manually annotated using Jalview v2.11.4.1 [59] following multiple sequence alignment with Clustal Omega [60]. Sequence alignments are available in the S1 File. A representative subset of 59 sequences (24 *Viannia*, 35 *Leishmania*) is illustrated in S1 Fig to indicate sequence conservation or variations across the two analyzed *Leishmania* subgenera. As for the 18S rDNA, 37 sequences from *Leishmania* spp. and 272 from *Trypanosoma* spp. were aligned and analyzed using the same bioinformatics pipeline (S2 File). A subset of 31 representative sequences (16 *Leishmania* spp., 15 *Trypanosoma* spp.) was selected to denote sequence conservation or variations across the two analyzed trypanosomatid genera (S2 Fig).

### crRNA guide sequences and preparation

We used previously reported LbCas12a crRNA guide sequences targeting 18S rDNA and kDNA minicircle sequences [45]. crRNA preparation from double-stranded DNA templates (see S1 Table for nucleotide sequences) was performed as described previously [45].

### Loop-mediated isothermal amplification (LAMP) assay

We first optimized the LAMP assay reaction conditions using a commercial Bst 2.0^®^ DNA polymerase (New England Biolabs Inc (NEB), Massachusetts, USA; Cat no. M0537), following the LAMP protocol as suggested by the manufacturer (NEB). The only modification was the final total concentration of MgSO_4_ in the reaction (6 mM Mg^2+^ for 18S rDNA and 5 mM Mg^2+^ for kDNA). These reaction conditions were used to evaluate the incubation time needed for optimal amplification of the target DNA, which was tested at 2 × 10^2^, 2 × 10^-1^, and 2 × 10^-3^ parasite genome equivalents (GE) per reaction to cover a wide range of target abundance. The LAMP reaction products were analyzed by 2% agarose gel electrophoresis using SYBR Gold staining and tested downstream using CRISPR-Cas12a assays with fluorescence readout (see below).

To facilitate master mix preparation for sample processing, LAMP reactions for the preamplification of *Leishmania* DNA targets were performed using the WarmStart® LAMP Kit (DNA & RNA) (NEB; Cat no. E1700). Each reaction contained 12.5 µL of WarmStart LAMP 2X Master Mix, 8 µL of molecular biology grade water, and 2.5 µL of 10X LAMP primer mix for a single target (kDNA or 18S rDNA). Like for the LAMP protocol using the separate Bst enzyme, the six target-specific primers (3 sets) were added at a final concentration of 1.6 µM for forward inner and backward inner primers (FIP/BIP), 0.4 µM for loop forward and loop backward primers (LF/LB), and 0.2 µM for forward external and backward external primers (F3/B3). After that, we carefully added 20 µL of immersion oil over the LAMP mix to create two layers to prevent cross contamination. Then, 2 µL of DNA template (0.26 – 40 ng, for analytical testing) was added to the first (bottom) layer, corresponding to the LAMP mixture, bringing the final reaction volume to 25 µL. The LAMP reaction was incubated on a heating block at 65°C for 30 min for kDNA or for 50 min for 18S rDNA, and then stopped by incubating at 82°C for 3 min.

A positive control (*L*. *braziliensis* M2904 gDNA, 10^2^ – 10^4^ GE as indicated in the figure legends) and a negative amplification control (LAMP reaction without DNA template, i.e., No-Template Control (NTC)) were included in all experiments. In the analytical specificity testing, a negative control consisting of human gDNA from peripheral blood mononuclear cells (PBMC) of a healthy donor (40LJng of input DNA) was also included.

### CRISPR-Cas12a assay with fluorescence readout

LbCas12a-based detection reactions were performed as described previously [45, 61, 62], with the only modification being that a different reporter probe was used, which allows fluorescence signal visualization on a LED transilluminator. We prepared the ribonucleoprotein (RNP) complex 10X by mixing recombinant LbCas12a (produced in-house [63]) with the target-specific folded crRNA (incubated at 65°C for 10 min followed by room temperature (RT) incubation for 10 min) in a 1:1.5 molar ratio and 2 µM single-stranded DNA (ssDNA) reporter probe (/56-FAM/TTATT/3IABkFQ/; [24]) in reaction buffer 1X (NEBuffer 2.1 recipe, prepared in-house, without adding MgCl_2_). The labeled fluorophore/quencher probe was purchased from Macrogen Inc. (Seoul, South Korea). A volume of 10 µL of the 10X RNP complex (pre-incubated for 10 min at RT) was loaded into each well of a flat-bottom, black 96-well microplate (Thermo Fisher Scientific, Waltham, MA, United States; Cat. no. 237107). Subsequently, 90 µL of the diluted LAMP product –prepared by diluting 2 µL of the LAMP product in 106 µL of 1X reaction buffer containing 16.7 mM MgCl_2_– was added. The 100 µL LbCas12a *trans*-cleavage reaction, with a final MgCl_2_ concentration of 15 mM, was incubated and analyzed in a fluorescence plate reader (Synergy™ H1 hybrid multi-mode reader; BioTek Instruments, Winooski, VT, United States) for 121 min at 25°C. Fluorescence measurements were recorded every 1 min (λex: 490 nm; λem: 525 nm) with a fluorescence gain setting of 120. For the experiments shown in S3 Fig, measurements used the Varioskan™ LUX multimode microplate reader (Thermo Fisher Scientific).

### Optimization of the CRISPR-Cas12a-based detection assay coupled with lateral flow readout

The lateral flow assay (LFA) readout was optimized using the HybriDetect - Universal lateral flow assay kit (Milenia Biotec GmbH, Giessen, Germany; Cat. no. MGHD 1) according to the manufacturer’s instructions. First, to optimize the reporter probe concentration (for reducing the background signal on the T-line of the LFA strip in negative controls), we tested the reporter at a final concentration (1X) of 500 nM, 250 nM, and 100 nM per LFA. Second, we tested the reaction buffer 1X without MgCl_2_ (prepared in-house following the NEBuffer 2.1 recipe) as compared to the HybriDetect universal assay buffer. Third, we tested two incubation temperatures of the Cas12a reaction: either 25°C or 37°C, for a reaction time of 30 min or 60 min. Fourth, we evaluated the addition of polyethylene glycol (PEG)-8000 at 2% and 4% final concentration to the completed CRISPR-Cas12a reaction for increasing the viscosity of the buffer, which in turn slows down lateral flow speed.

### Optimized CRISPR-Cas12a-based lateral flow strip assay for clinical samples and reference strain genomic DNA

After completion of the preamplification step, 2 µL of the LAMP product was mixed with 106 µL of 1X reaction buffer with 16.7 mM MgCl_2_ in a 1.5 mL microcentrifuge tube. Then, we took 90 µL of the diluted LAMP product in a new microcentrifuge tube and added 10 µL of the 10X pre-incubated RNP complex (100 nM Cas12a: 150 nM crRNA: 1000 nM reporter probe). The lateral flow cleavage reporter probe (5’/Bio/TTATTATT/6-FAM/3’; with terminal biotin and FAM labels in the opposite order relative to [26]) was purchased as an HPLC-purified oligo from biomers.net GmbH (Ulm, Germany). The 100 µL LbCas12a *trans*-cleavage reaction, with a final MgCl_2_ concentration of 15 mM, was incubated at 37°C for 60 min in the dark. Afterward, 5 µL of PEG-8000 (2% final concentration) was added to the reaction tube and homogenized by pipetting up and down. A lateral flow strip (Milenia HybriDetect) was placed into the reaction tube and incubated for 5 min at RT. Lateral flow strips were interpreted visually. LFA strip images shown here were taken with an iPhone 13 Pro camera.

### Analytical sensitivity testing

Serial dilutions of *L*. *braziliensis* M2904 gDNA (extracted from a promastigote culture) to a final range of 2 × 10^4^ to 2 × 10^−3^ parasite GE per reaction (encompassing the same range of parasite GE per reaction of the standard curve used in the kDNA qPCR assay [64]) were tested as input DNA in LAMP reactions to amplify the target gene, followed by LbCas12a-based detection assays as described above, using a plate-based fluorescence readout and a colorimetric lateral flow-based readout. The analytical sensitivity for each target gene was determined based on 3 independent experiments.

### Analytical specificity testing

The analytical specificity of the LAMP-CRISPR assays targeting *Leishmania* kDNA (*Viannia* subgenus) or 18S rDNA was tested using gDNA from representative laboratory reference strains of New World and Old World *Leishmania* species, as well as other microbial pathogens (Table 1). Target genes were amplified by LAMP (using 20–40 ng of input DNA) and detected by LbCas12a-based detection assays as described above, using fluorescence-based and lateral flow-based readouts. Two independent LAMP-CRISPR experiments were performed.

### Evaluation of LAMP-CRISPR assay performance on clinical samples

Patient DNA samples extracted from skin lesion specimens were tested blindly by the LAMP-CRISPR assays in groups of ten, plus positive and negative controls. The DNA input for the LAMP preamplification step was 5 µL of diluted or undiluted DNA samples (6–285 ng of input DNA; see S1 Dataset) to capture data from samples with low *Leishmania* parasite load levels. The LAMP products were then detected by LbCas12a-based detection assays as described above.

### Quantitative real-time PCR (qPCR)

A qPCR assay targeting *Leishmania* kDNA minicircles was performed to detect and quantify *Leishmania* (*Viannia*) parasites in clinical samples, using primers and cycling conditions as described [64]. The human *ERV-3* gene was amplified in parallel using the primers reported by Yuan *et al*. [65]; it served as an indicator of specimen adequacy and to estimate the parasite load normalized to the number of human cell equivalents per sample [64].

### Data processing and analysis

The data collected from LAMP-CRISPR assays were analyzed as described previously [45]. Briefly, the raw fluorescence data from each well of the LbCas12a-based assay plate were exported to Microsoft Excel. Based on the inspection of fluorescence time-course data from samples and controls that were run in parallel, the time point of fluorescence accumulation for data analysis was defined (at 25-min time point for both kDNA and 18S rDNA). The raw fluorescence data were normalized by dividing target reaction fluorescence accumulated at the defined time point of the test sample to that of the NTC reaction included in parallel in the same Cas12a assay plate (further called ‘fluorescence ratio’). In the analytical validation, a result was considered ‘detected’ if a target reaction produced a fluorescence ratio ≥ 2 for the test sample above background (NTC). To set a threshold cutoff value for positive signal (i.e., detection of *Leishmania*) in clinical samples, we required that the fluorescence ratio be larger than the mean and three standard deviations of negative clinical samples (see S1 Dataset) selected as a separate panel for determining the cutoff. Graphs, numerical data analyses, and statistical analyses were performed using GraphPad Prism version 10.2 (GraphPad Software, San Diego, CA, United States).

For the diagnostic performance evaluation of the LAMP-CRISPR assays with fluorescence readout, the sensitivity and specificity and their 95% confidence intervals were calculated with MedCalc software [66] using kDNA real-time PCR as the reference test. To evaluate the accuracy of the LAMP-CRISPR assays to discriminate a positive (detected) from a negative (not detected) *Leishmania* infection status, a receiver operating characteristic (ROC) curve was drawn and the area under the curve (AUC) with its 95% confidence interval was calculated using GraphPad Prism version 10.2. In all statistical tests, a *P* value ofLJ<LJ0.05 was considered statistically significant.

To assess the applicability of the LAMP-CRISPR assays with an LFA readout, 18 clinical samples selected for pilot testing were arbitrarily categorized into three groups based on parasite load: low (<100 parasites/10^6^ human cells), intermediate (100 – 10,000 parasites/10^6^ human cells), and high (>10,000 parasites/10^6^ human cells). LFA strips were interpreted blinded to the fluorescence results.

## Results

### Design and optimization of LAMP-CRISPR assays

We developed two assays for the detection of *Leishmania* spp., which are based on the most frequently used multicopy genetic markers in molecular diagnostics for leishmaniasis: kDNA minicircles and 18S rDNA. Our assays combine LAMP with CRISPR-LbCas12a-mediated DNA target recognition and ssDNA reporter cleavage, with results assessed either with a fluorescence-based or lateral flow strip readout (Fig 1). These two readouts are made possible through the use of different dual-labeled ssDNA reporter molecules: one labeled with a fluorophore-quencher pair for real-time monitoring of fluorescence curves over the Cas12a reaction time course on a plate reader, and the other labeled with biotin and FAM for results to be visually read with paper-based lateral flow strips [26].

**Fig 1.**
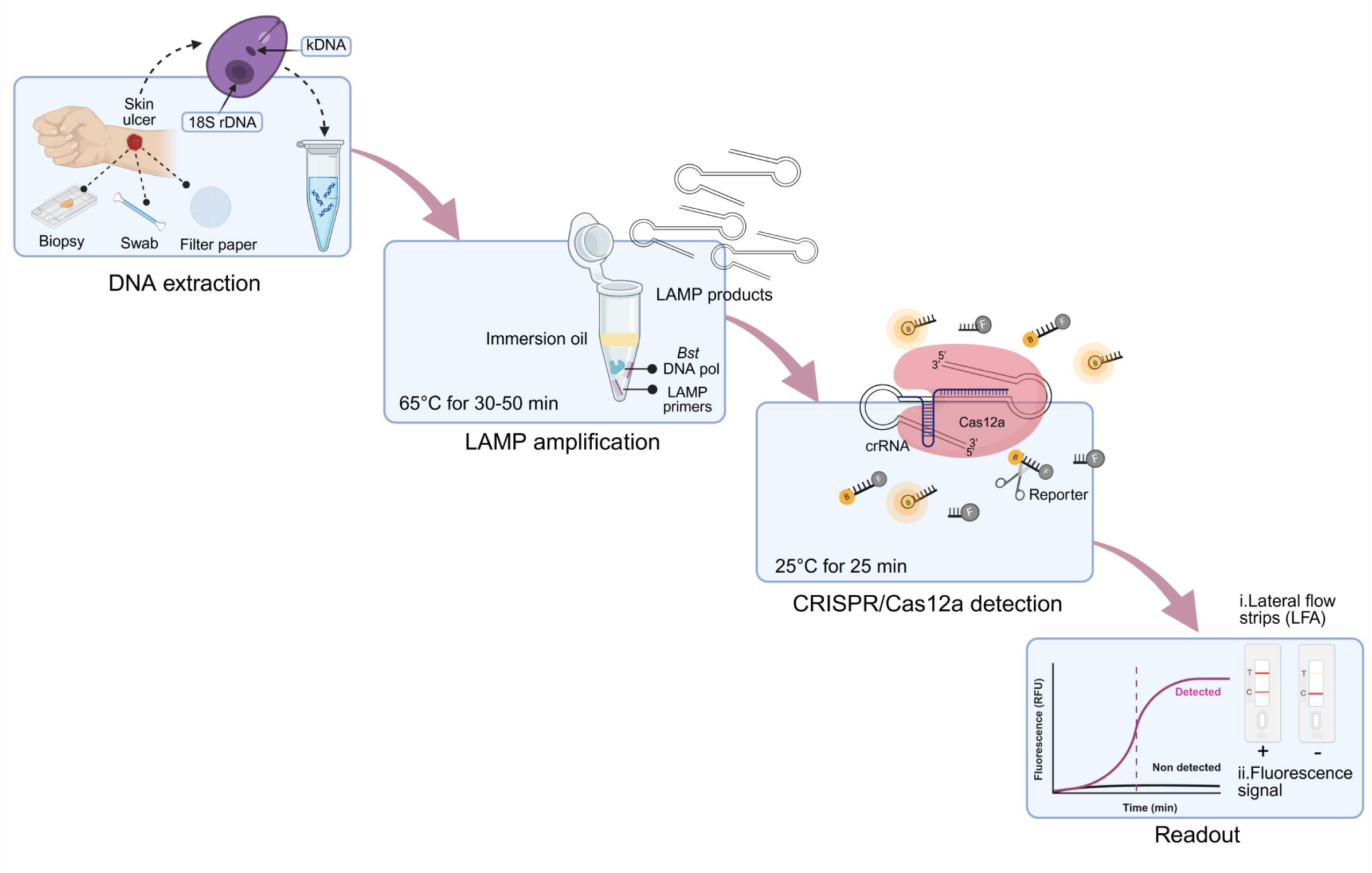
Schematic representation of the workflow of LAMP-CRISPR assays for detection of *Leishmania* spp. in clinical samples. Sample preparation is performed from skin lesion specimens taken from patients with suspected CL. The DNA samples are subjected to LAMP amplification for just 30 min (kDNA) or 50 min (18S rDNA), followed by Cas12a-based reactions with either a fluorescence-based or lateral flow strip readout. Figure created with BioRender.com.

We designed LAMP primers flanking previously reported crRNA target recognition sites [45] within *Leishmania* kDNA minicircles (S1 Fig) and 18S rDNA (S2 Fig). According to the *in silico* analyses, the kDNA primer set targets a region most conserved within the *L*. (*Viannia*) subgenus and less conserved within the *L*. (*Leishmania*) subgenus (S1 Fig and S1 File). *In silico* analysis of the selected 18S primer set showed the expected conserved nature of the targeted genomic region at the *Leishmania* genus level, whereas sequence variations were more frequent within the *Trypanosoma* genus (S2 Fig and S2 File).

A schematic depicting the mechanism of LAMP amplification, with the resulting LAMP amplicon containing the Cas12a crRNA target sequence is shown in Fig 2. We optimized the LAMP assay reaction conditions using a commercial Bst 2.0 DNA polymerase and tested the incubation time required for optimal amplification of the target DNA. The CRISPR-Cas12a assay was used as the readout. For kDNA, the LAMP reaction was evaluated at 4 different incubation times (10, 20, 30, and 40 min) (S3A-D Fig). While 10 min were sufficient to amplify the target DNA sequence present at a high quantity (2 × 10^2^ GE) (S3A Fig), at least 20 min were required for the amplification of low abundant target DNA (2 × 10^-1^ GE) (S3B Fig). On the other hand, an incubation time of 40 min enabled the amplification of a very low quantity of the target DNA (2 × 10^-3^ GE) but also resulted in a positive signal of the NTC reaction (S3D Fig), the latter indicative of contamination. These results were corroborated through visualization of the LAMP reaction products by agarose gel electrophoresis (S3F Fig). We thus chose an incubation time of 30 min as the most optimal for the kDNA LAMP reaction (S3C and S3E Fig) for further experiments. For the 18S rDNA gene, we tested the LAMP reaction at 5 different incubation times (10, 30, 40, 50, and 60 min) (S3G-K Fig). At least 40 min were required to amplify the target DNA present at varying levels of abundance (2 × 10^2^ and 2 × 10^-1^ GE) (S3I Fig). Extending the incubation time of the LAMP reaction to 60 min allowed the amplification of very low-abundant target DNA (2 × 10^-3^ GE) but also resulted in NTC contamination (1 out of 2 replicates of the NTC reaction showed a positive signal) (S3K Fig). CRISPR-Cas12a-based detection of LAMP amplicons could effectively discriminate between specific amplification products and non-specific amplification. The latter was observed, for instance, at the incubation times of 10 min and 40 min in the NTC reaction by agarose gel electrophoresis (S3M Fig) but did not result in false positives in the Cas12a assay (S3G and S3I Fig). On the basis of these results, we selected an incubation time of 50 min for the 18S LAMP reaction (S3J and S3L Fig) for further experiments.

**Fig 2.**
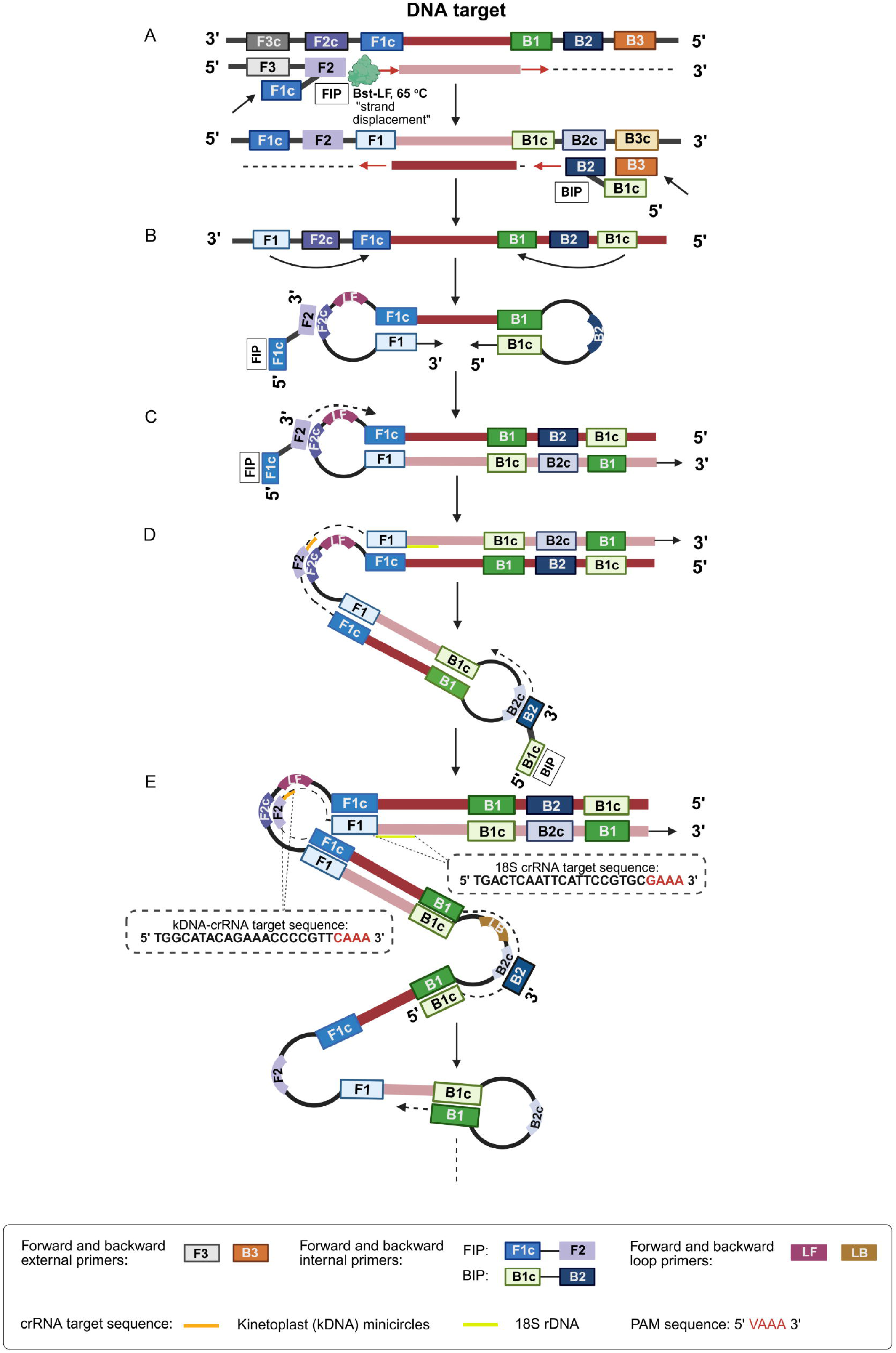
Schematic representation of the LAMP reaction, highlighting the location of the crRNA target sequence in the target DNA. (A-E) Steps in the LAMP reaction to amplify the target DNA. The eight annealing locations of the six LAMP primers are shown. The primers were designed to target unique sequences within a conserved region of *Leishmania* kDNA minicircles (S1 Fig) or nuclear 18S rDNA (S2 Fig). The crRNA target sequence and protospacer adjacent motif (PAM) site in the target DNA are shown. For simplicity, the locations of both crRNA target sites are depicted in the same graph (actually, the LAMP reaction is performed separately for each target). Figure adapted from [14, 67] and created with BioRender.com.

The CRISPR-Cas12a assay is based on purified recombinant LbCas12a [63]. For the fluorescence-based assay, we used optimized conditions as described previously [45, 61, 62] and a quenched fluorescent ssDNA reporter [24]. We optimized here the performance of the Cas12a-based LFA readout by testing varying concentrations of the biotin-FAM reporter probe as well as the influence of the assay buffer, incubation temperature and reaction time of the Cas12a assay, and PEG-8000 addition. The optimal conditions identified involved: using the reporter probe at a final concentration (1X) of 100 nM in our in-house Cas12a reaction buffer 1X (which performed comparable to the HybriDetect universal assay buffer); incubating the Cas12a reaction at 37°C for 60 min; and adding 2% PEG-8000 to the completed Cas12a reaction (S4 Fig).

### Analytical validation of LAMP-CRISPR assays

We then assessed the analytical sensitivity of each primer set-crRNA combination using serial dilutions of *L. braziliensis* M2904 gDNA extracted from a promastigote culture. The kDNA LAMP-CRISPR assay displayed positive *L. braziliensis* detection down to 2 × 10^-1^ GE per reaction, both with a fluorescence readout (Fig 3A) and a LFA readout (Fig 3B). Visualization of the kDNA LAMP products on an agarose gel confirmed the presence of ladder-like bands indicative of a successful LAMP reaction (Fig 3C), consistent with the Cas12a-based detected amplicon samples (Fig 3A and 3B). We found a similar analytical sensitivity for the 18S LAMP-CRISPR assay, as detection of at least 2 × 10^-1^ GE/reaction was achieved with a fluorescence-based readout (Fig 3D) and a LFA readout (Fig 3E). The analysis of the 18S LAMP products by agarose gel electrophoresis (Fig 3F) was consistent with the Cas12a assay results (Fig 3D and 3E). The no-template control (NTC) tested in parallel in each LAMP-CRISPR assay showed a background signal in the Cas12a fluorescence-based assay (Fig 3A and 3D, see the two lowest GE input amounts whose fluorescence ratio is 1 given that the denominator is the NTC signal). Likewise, the NTC did not exhibit a band (or the band observed was faint) at the test line in the lateral flow strips (Fig 3B and 3E). This ruled out the possibility of cross-contamination and non-specific amplification.

**Fig 3.**
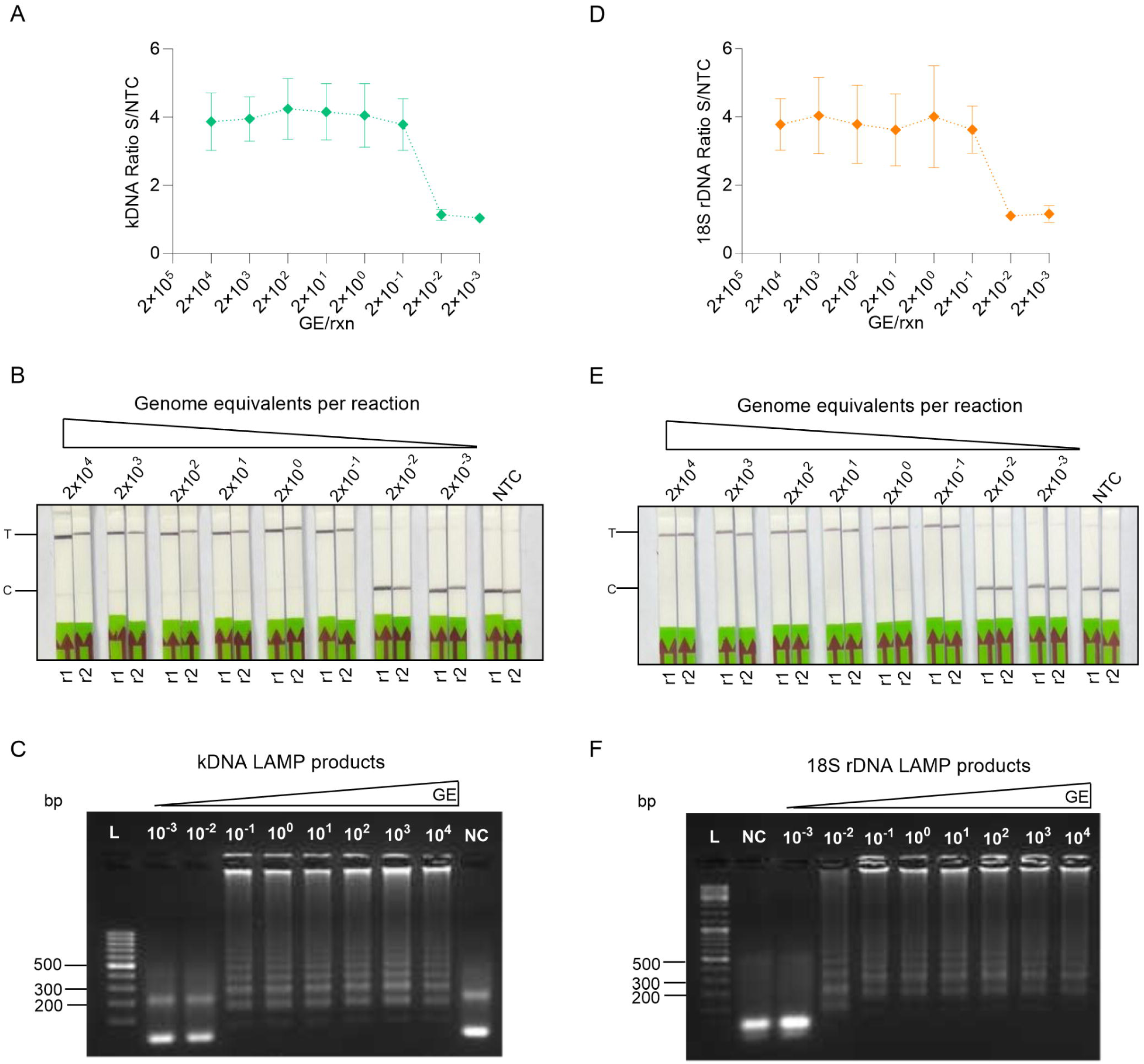
Analytical sensitivity testing of kDNA and 18S LAMP-CRISPR assays. Serial dilutions of *L. braziliensis* M2904 gDNA encompassing 2 × 10^4^ to 2 × 10^-3^ genome equivalents (GE) per reaction were subjected to LAMP amplification, followed by Cas12a-based reactions for the kDNA (A-C) or 18S rDNA (D-F) target. (A, D) Normalized fluorescence signals (fluorescence ratio) from Cas12a reactions (i.e., fluorescence signals taken at *t* = 25 min in the test sample relative to the NTC).

Fluorescence measurements were taken on the Synergy H1 plate reader. Data are represented as mean ± SD (n = 3 independent amplification and detection runs). The X-axis has a logarithmic scale. A fluorescence ratio ≥ 2 is considered detected. (B, E) Cas12a reactions were performed using a biotin-FAM reporter molecule and visualized on lateral flow strips. Uncleaved reporter molecules are captured at the first detection line (control line), whereas collateral Cas12a cleavage activity generates a signal at the second detection line (test line). The arrow indicates the direction of flow. Two independent amplification and detection runs (r1 and r2) were analyzed. (C, F) LAMP reaction products (5 µL) visualized by 2% agarose gel electrophoresis ran at 100 V for 1 h using SYBR Gold staining. L, GeneRuler 100 bp (C) or 100 bp Plus (F) DNA ladder. NC, negative control (NTC, no-template control). Since space was tight on the agarose gel images, we had to abbreviate the labels for each lane. For instance, “10^4^” actually represents “2 × 10^4^” as indicated in the fluorescence (A, D) and LFA (B, E) panels.

Assay specificity testing (S5 Fig) was performed using gDNA samples isolated from cultured laboratory reference strains of *Leishmania* spp., phylogenetically close pathogens, and pathogens that cause similar lesions to CL or can coinfect the same host (Table 1). The kDNA LAMP-CRISPR assay consistently detected representative strains of the *Leishmania* (*Viannia*) species present in our sample set (10 of 10; Fig 4A). This was expected given that our designed kDNA-crRNA targets a region of kDNA minicircles conserved within the *L*. (*Viannia*) subgenus ([45] and S1 Fig). Additionally, the assay detected 2 out of 6 tested strains belonging to the *L*. (*Leishmania*) subgenus (Fig 4A). The 18S LAMP-CRISPR assay showed pan-*Leishmania* detection ability (Fig 4A). In both kDNA and 18S LAMP-CRISPR assays, no cross-reactivity was observed for related trypanosomatid (*T. cruzi*) strains and other protozoan (*P. falciparum*) and bacterial (*M. tuberculosis*) pathogens tested (Fig 4A). Also, the negative control with human gDNA as template was not detected (Fig 4A). The analytical specificity was also assessed with an LFA readout, where strips were interpreted by visual inspection on the basis of the presence or absence of test bands. Three *L*. (*Viannia*) strains used as positive controls were detected (i.e., showing a strong T-line and a very weak or missing C-line) by both LAMP-CRISPR assays (Fig 4B, strips # 3-5). The negative controls (human gDNA and NTC) had a negative test result (Fig 4B, strips # 1 and 2). There was no cross-reactivity to other microbial pathogens tested (Fig 4B, strips # 6-10, which show a strong C-line and a faint (or absent) T-line, similar to the negative controls).

**Fig 4.**
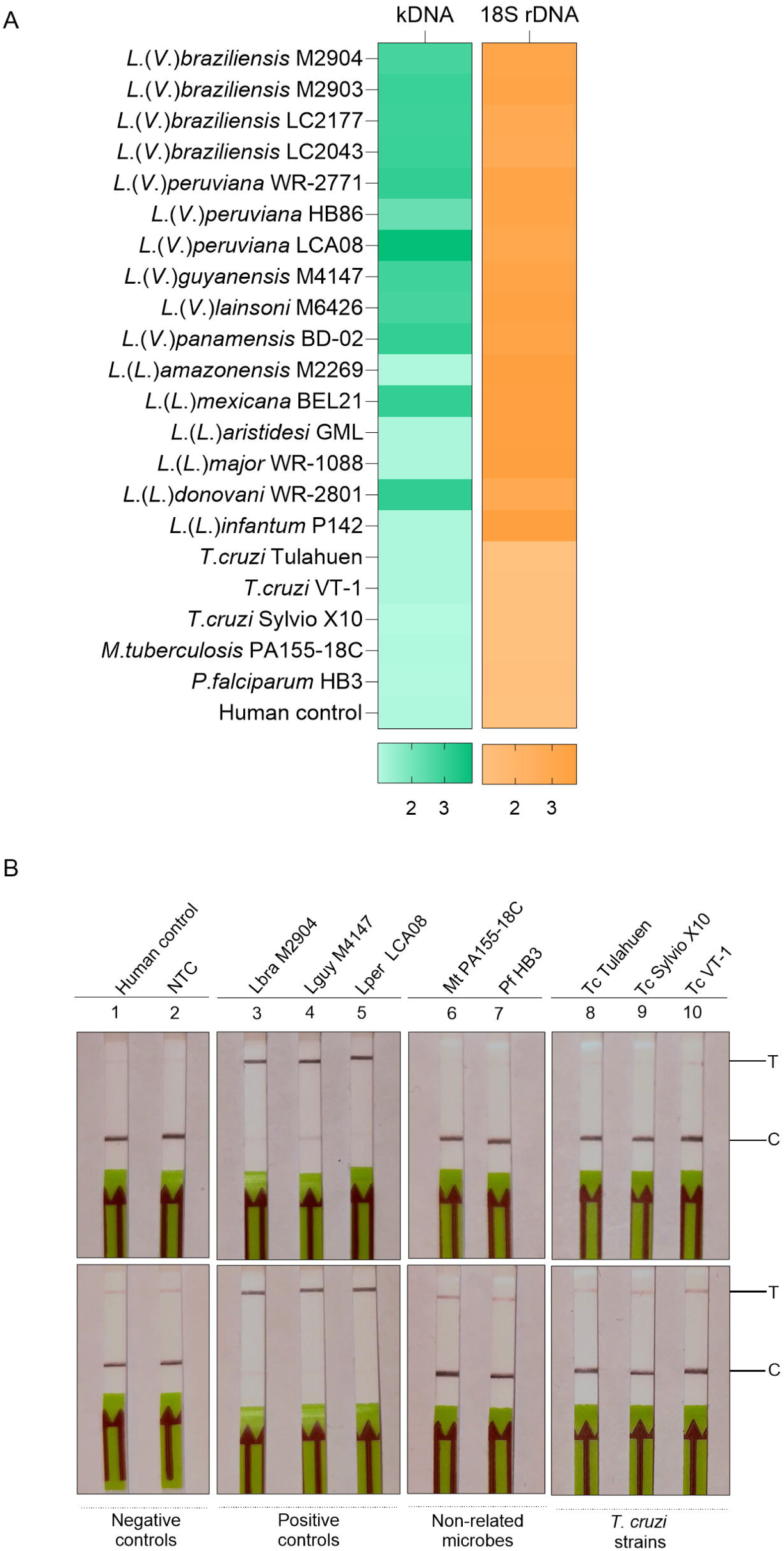
Analytical specificity testing of kDNA and 18S LAMP-CRISPR assays. The specificity of the kDNA and 18S LAMP-CRISPR assays was evaluated using gDNA from reference strains of *Leishmania* spp. and other microbial pathogens (see Table 1 and S5 Fig). (A) Heat maps depicting LAMP-CRISPR fluorescence-based detection data from tested reference strain samples. The color scale represents normalized fluorescence signals from Cas12a reactions (i.e., fluorescence signals taken at *t* = 25 min in the test sample relative to the NTC) for *Leishmania* kDNA and 18S rDNA targets. The fluorescence ratio values from 2 independent assay repeats were entered into GraphPad; the mean of these replicates is displayed. A fluorescence ratio ≥ 2 is considered detected. (B) Cas12a reactions were performed using a biotin-FAM reporter molecule and visualized on lateral flow strips. Uncleaved reporter molecules are captured at the first detection line (control line), whereas collateral Cas12a cleavage activity generates a signal at the second detection line (test line). The arrow indicates the direction of flow. Top panel: kDNA LAMP-CRISPR assay; bottom panel: 18S LAMP-CRISPR assay. Negative controls: HC, human control (gDNA from PBMCs of a healthy donor) (strip # 1); NC, no-template control (NTC) (strip # 2). Positive controls: Lbra, *L. braziliensis* M2904 (strip # 3); Lguy, *L. guyanensis* M4147 (strip # 4); Lper, *L. peruviana* LCA08 (strip # 5). Non-related microbes: Mtb, *M. tuberculosis* PA155-18C (strip # 6); Pf, *P. falciparum* HB3 (strip # 7). *T. cruzi* (Tc) strains: Tulahuen, Sylvio X10, and VT-1 (strips # 8-10).

### Performance evaluation of LAMP-CRISPR assays with clinical samples

Positive patient samples exhibited robust fluorescence curves in the Cas12a assay indicating the presence of *Leishmania* kDNA (S6A Fig) or 18S rDNA (S6B Fig) molecules. These results were consistent with the respective positive LAMP reactions as visualized on an agarose gel (S6C and S6D Fig). We noted that the analysis of LAMP products by agarose gel electrophoresis can sometimes give ambiguous (or difficult to interpret) results. For instance, ladder-like bands were observed for samples S72 and S09 amplified by the 18S LAMP assay (S6D Fig), although the band pattern was different from the one observed for true positive reactions. This was clearly resolved by the Cas12a-based assay on those amplicons, which confirmed that S72 and S09 were negative for *Leishmania* 18S rDNA molecules (S6B Fig).

We next evaluated the performance of our LAMP-CRISPR fluorescence-based detection assays using gDNA from 90 clinical samples of skin lesions. An in-house qPCR assay [64] targeting a kDNA minicircle region conserved among *Leishmania* (*Viannia*) species [68] served as the reference test. Of the 90 samples, 55 were *Leishmania*-positive and 35 were negative by qPCR.

To determine the cutoff value for discriminating between *Leishmania*-positive and –negative samples by the LAMP-CRISPR assays, we used the mean of the fluorescence ratio of negative samples + 3SD. To this end, an independent panel of negative samples was used: n = 19 for the kDNA assay and n = 10 for the 18S assay (S1 Dataset). A higher number of negative samples was needed for the former to capture the inter-assay variation in raw fluorescence signal from the Cas12a reaction that was evident over different periods of sample testing (S1 Dataset). The determined cutoff value was 1.46 for the kDNA assay and 1.30 for the 18S assay (S1 Dataset).

The kDNA LAMP-CRISPR assay detected 50 (90.9%) of 55 qPCR-positive samples (Fig 5 and 6A, Table 2, and S1 Dataset). Of these, the 5 samples not detected by kDNA LAMP-CRISPR (coded 3, 5, 6, 28 and 71) harbored low parasite loads (≤20 parasites/10^6^ human cells) (Fig 6B and S1 Dataset), corresponding to 10^-2^ to 10^-3^ parasite GE/reaction (S1 Dataset, see “Parasite equivalents” data in the worksheet “qPCR_CS”). The 18S LAMP-CRISPR assay detected 40 (72.7%) of 55 qPCR-positive samples (Fig 5 and 6C, Table 2, and S1 Dataset). Of these, the 15 samples not detected by 18S LAMP-CRISPR contained a parasite load of ≤150 parasites/10^6^ human cells (Fig 6D and S1 Dataset), which corresponded to 10^-1^ to 10^-3^ parasite GE/reaction (S1 Dataset, see “Parasite equivalents” data in the worksheet “qPCR_CS”). Both LAMP-CRISPR assays achieved 100% specificity (Table 2). The complete dataset of this study is presented in the S1 Dataset.

**Fig 5.**
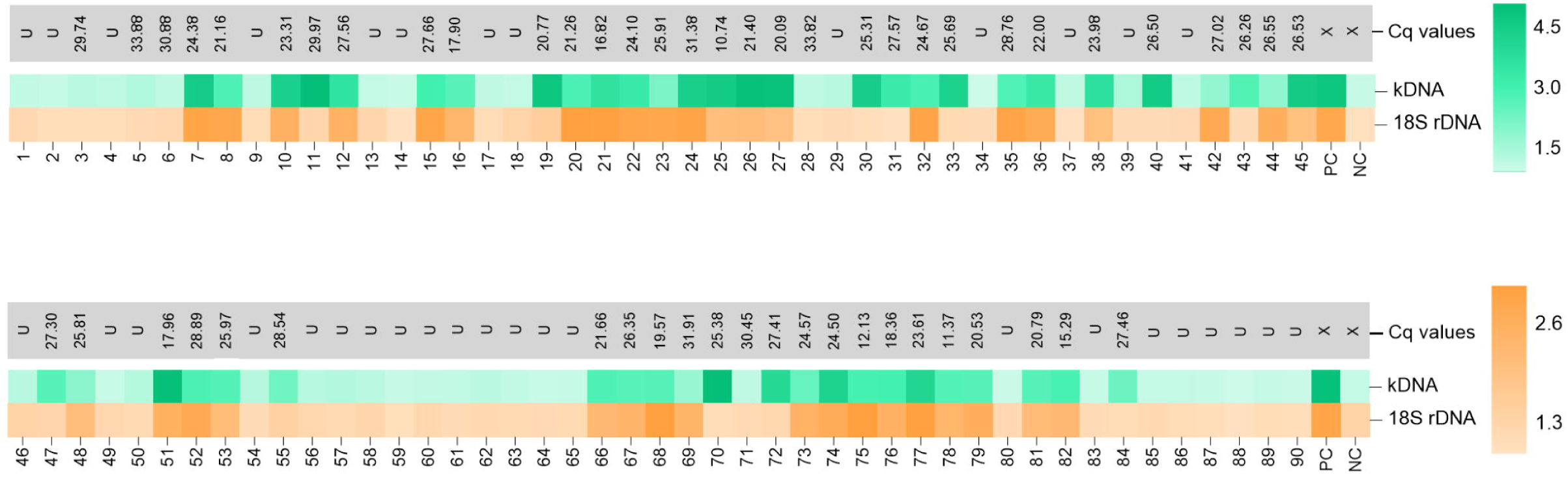
Evaluation of kDNA and 18S LAMP-CRISPR assay performance on clinical samples. Heat maps showing LAMP-CRISPR fluorescence-based detection data from tested clinical samples (n = 90). The color scale represents normalized fluorescence signals from Cas12a reactions (i.e., fluorescence signals taken at *t* = 25 min in the test sample relative to the NTC) for *Leishmania* kDNA and 18S rDNA targets. Samples are coded as 1-90 (‘Code_UPC’ as indicated in the S1 Dataset). PC, positive control (*L. braziliensis* M2904 gDNA, 10^3^ – 10^4^ genome equivalents); NC, no-template control (NTC) of the LAMP reaction. The cutoff value was 1.46 for the kDNA assay and 1.30 for the 18S assay (mean of the fluorescence ratio of negative clinical samples used to calculate cutoff values + 3SD; see the S1 Dataset). The Cq values corresponding to the kDNA qPCR assay (reference test) are shown. For Cq values, the letter “U” denotes undetermined; X, not applicable.

**Fig 6.**
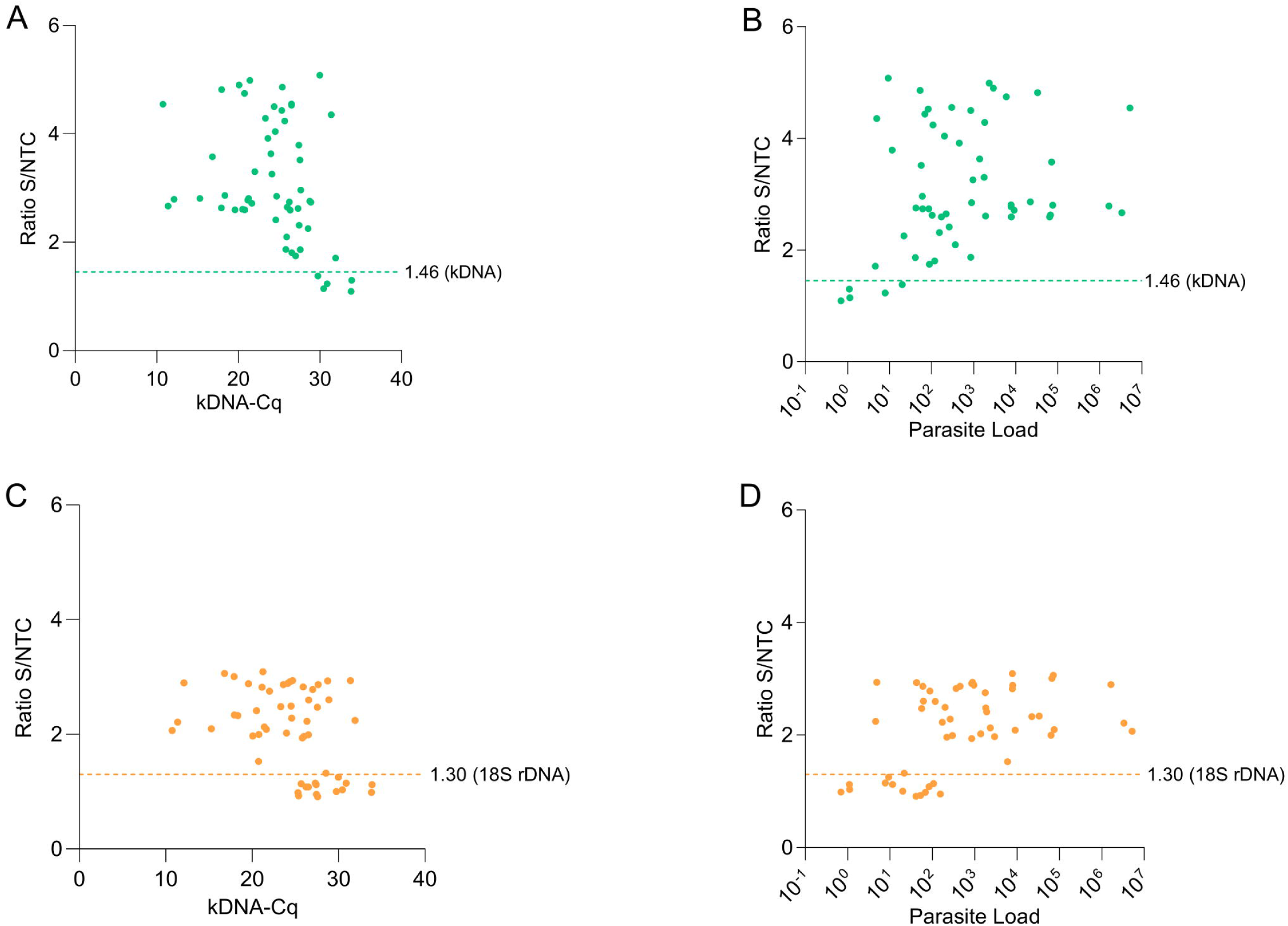
Evaluation of kDNA and 18S LAMP-CRISPR assay performance on clinical samples: analysis across the range of parasite load levels. (A, C) Scatter plots showing normalized fluorescence signals from Cas12a reactions (i.e., fluorescence signals taken at *t* = 25 min in the test sample relative to the NTC) for *Leishmania* kDNA (A) and 18S rDNA (C) targets in clinical samples (n = 90; data shown in Fig 5 and S1 Dataset) versus Cq values determined by kDNA qPCR. (B, D) Scatter plots showing normalized fluorescence signals from Cas12a reactions for *Leishmania* kDNA (B) and 18S rDNA (D) targets in clinical samples versus the estimated parasite load in quantifiable samples (n = 55). The parasite load is expressed as the number of *Leishmania* parasites/10^6^ human cells. The horizontal dotted lines represent the LAMP-CRISPR assay cutoff value (mean of the fluorescence ratio of negative clinical samples + 3SD; see the S1 Dataset): 1.46 for the kDNA assay and 1.30 for the 18S assay.

**Table 2.** Diagnostic performance of LAMP-CRISPR assays with fluorescence readout for *Leishmania* DNA detection compared with the kDNA qPCR assay.

| Test | kDNA qPCR |  | % (95% CI) |  |
| --- | --- | --- | --- | --- |
|  | Positive<br>(n = 55) | Negative<br>(n = 35) | Sensitivity | Specificity |
| kDNA LAMP-CRISPR |  |  | 90.91 (80.05-96.98) | 100.00 (90.00-100.00) |
| Positive | 50 | 0 |  |  |
| Negative | 5 | 35 |  |  |
| 18S LAMP-CRISPR |  |  | 72.73 (59.04-83.86) | 100.00 (90.00-100.00) |
| Positive | 40 | 0 |  |  |
| Negative | 15 | 35 |  |  |

Results of the statistical analysis of diagnostic test performance conducted with the MedCalc software version 23.0.1. (https://www.medcalc.org/calc/diagnostic_test.php)

A ROC curve analysis was used to assess the performance of each assay in distinguishing *Leishmania*-positive and -negative clinical samples, with AUC values of 0.973 for the kDNA LAMP-CRISPR assay (Fig 7A) and 0.876 for the 18S LAMP-CRISPR assay (Fig 7B).

**Fig 7.**
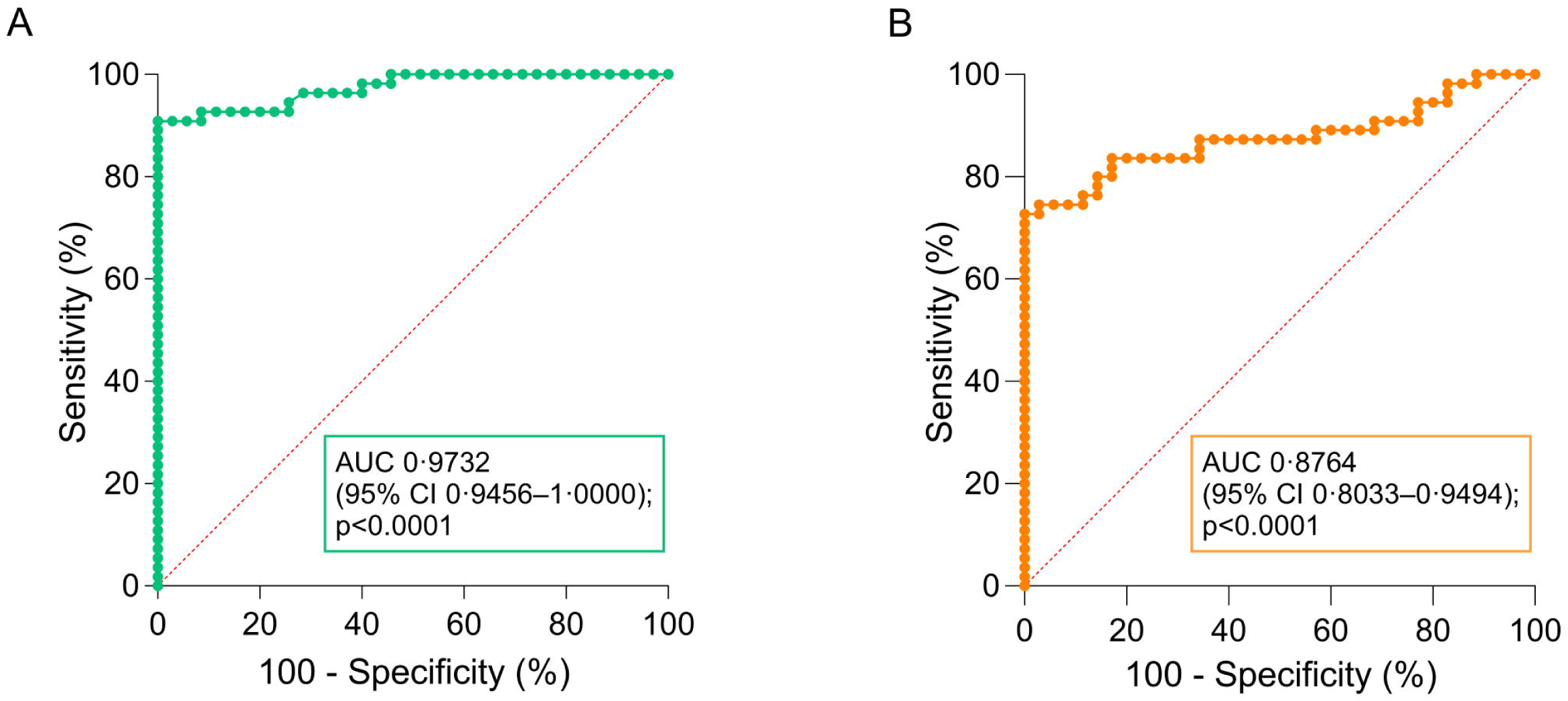
Evaluation of kDNA and 18S LAMP-CRISPR assay performance on clinical samples: ROC curve analysis. ROC curve analysis performed using the fluorescence ratio values of the kDNA LAMP-CRISPR assay (A) or the 18S LAMP-CRISPR assay (B) from tested clinical samples (n = 90; data shown in Fig 5 and S1 Dataset), with the *Leishmania* infection status (positive/negative) as determined by kDNA qPCR (reference test). The area under the curve (AUC) and corresponding 95% confidence interval (calculated using the Wilson/Brown method) are shown. The diagonal dotted line corresponds to the null hypothesis with AUC of 0.5, in which the accuracy of a diagnostic test is no different from random chance.

We also assessed the applicability of the LAMP-CRISPR assays with an LFA readout on clinical samples. To this end, we performed pilot testing on gDNA extracted from a representative group of clinical samples (n = 18) encompassing a wide range of parasite load levels as determined by the kDNA qPCR assay. Lateral flow testing results were consistent across two assay repeats per sample (n = 8; S7 Fig), indicating robust assay reproducibility. Subsequently, we expanded the evaluation to include 10 additional clinical samples, thus analyzing a total of 18 specimens (Fig 8). Samples that tested positive by the kDNA LAMP-CRISPR assay (16/18 = 88.9%) included the 3 parasite load categories: low (4 of 6 tested), intermediate (6 of 6), and high (6 of 6) (Fig 8). These LFA results were 100% concordant with the fluorescence-based readout (Fig 8 and S1 Dataset). The 18S LAMP-CRISPR assay with LFA readout detected *Leishmania* DNA in 72.2% (13/18) of the examined samples, which had parasite load levels defined as: low (2 of 6 tested), intermediate (5 of 6), and high (6 of 6) (Fig 8). One sample (coded 12) out of the 18 tested samples (5.6%) by the 18S assay showed discordant results between LFA and fluorescence-based readouts (Fig 8), with this associated with a low parasite load (56.6 parasites/10^6^ human cells) (S1 Dataset). The two examined DNA samples from non-leishmaniasis patients (i.e., kDNA qPCR-negative; samples 09 and C7) tested negative by both LAMP-CRISPR assays (Fig 8 and S7 Fig).

**Fig 8.**
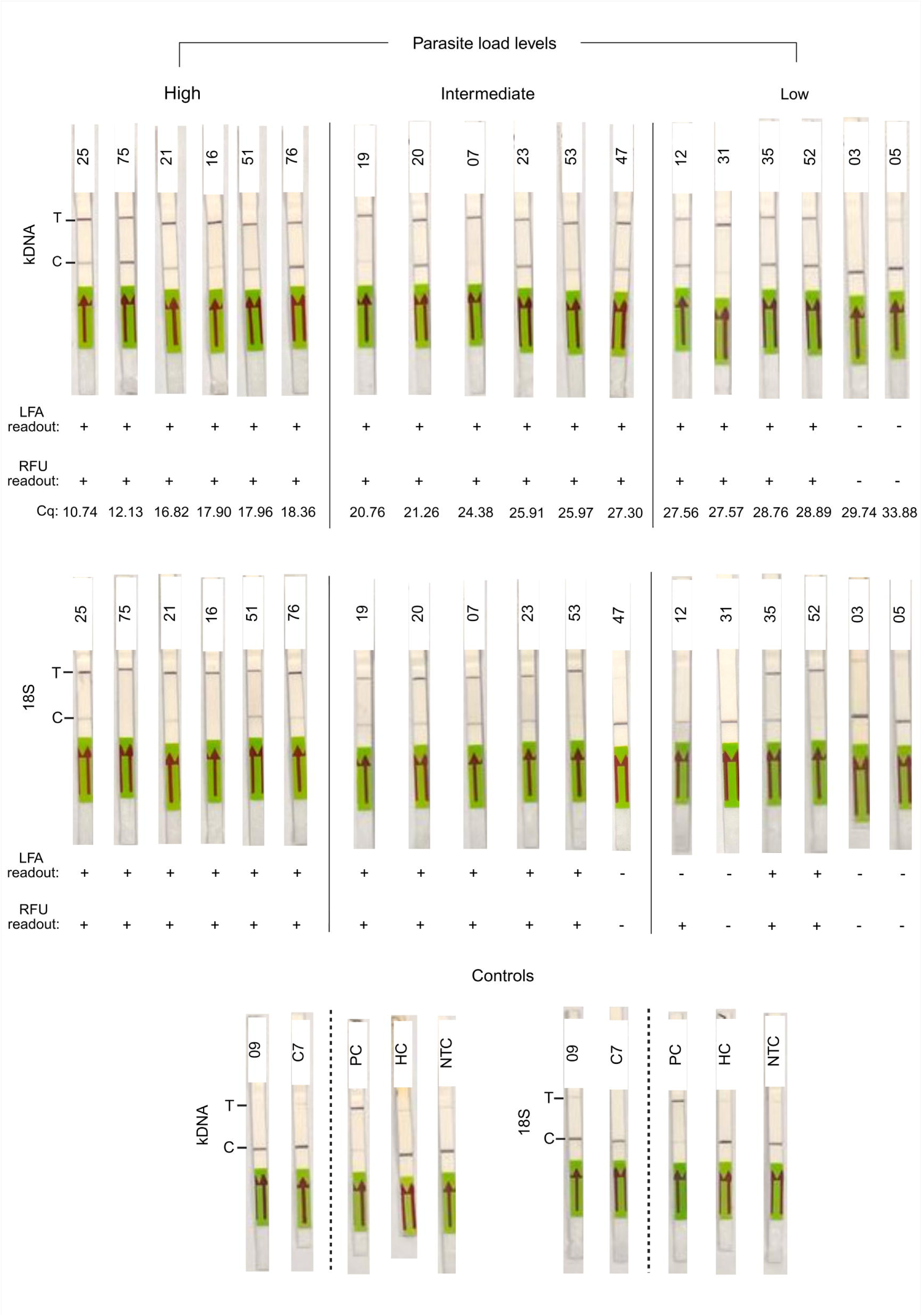
Pilot testing of kDNA and 18S LAMP-CRISPR assays on clinical samples using a lateral flow readout. A representative subset of clinical samples with varying parasite load levels was tested with kDNA and 18S LAMP-CRISPR assays using a LFA readout. Visualization on the lateral flow strips (Milenia GenLine HybriDetect strips) was achieved through the cleavage of the biotin-FAM reporter molecule. C, control line; T, test line. The results of the fluorescence-based readout of the LAMP-CRISPR assays (+, detected; -, not detected, on the basis of the estimated cutoff; see S1 Dataset) as well as the Cq values of the kDNA qPCR are indicated below each strip. Controls included DNA samples from two non-leishmaniasis patients (i.e., kDNA qPCR-negative; samples 09 and C7), a positive control (PC, 2 × 10^2^ genome equivalents from *L. braziliensis* M2904 gDNA), a human negative control (HC, 20 ng of human PBMC gDNA), and a no-template control (NTC) of the LAMP reaction.

## Discussion

The pressing need for new diagnostic tools has been identified as a priority in the World Health Organization (WHO) 2030 road map to support the control and elimination of neglected tropical diseases [69]. Among them, leishmaniasis belongs to the category of the most neglected ones as it entails a group of diseases with a large negative impact on people living in endemic rural areas, who are often economically disadvantaged and do not have access to an early, correct disease diagnosis that would enable timely adequate treatment. PCR-based molecular diagnostics provide the most sensitive and specific tools for *Leishmania* detection and are pivotal for early disease diagnosis, discrimination of *Leishmania* species, detection of asymptomatic infections, and epidemiological surveillance [70]. Besides their use in centralized laboratories, the progress in the automation and simplification of the workflow from sample preparation to target amplification and detection (result readout) steps has made molecular diagnostics more suitable and accessible for resource-limited settings in the form of near-PoC tests [71]. Examples of promising formats include the *Leishmania* OligoC-TesT (a PCR-oligochromatographic test) [72, 73] and a PCR-based method applied directly on EDTA blood for the diagnosis of VL, where the PCR is run on a portable miniPCR device and the labeled PCR products are detected with a lateral flow immunoassay [74]. Other recent advances have been facilitated by the incorporation of isothermal NAATs, such as LAMP and RPA, combined with lateral flow detection or other portable devices [75–79]. Among the most promising approaches is the integration of isothermal NAATs with CRISPR technology, which has fostered the development of a new generation of molecular diagnostics with the potential to advance toward near-PoC applications in low-resource settings [80].

In the present study, we aimed to contribute with the development of LAMP-coupled CRISPR-Cas12a assays for molecular detection of *Leishmania* spp. We designed new LAMP primer sets that flank previously reported crRNA target sites [45] within two commonly used multicopy regions of the *Leishmania* genome: the 18S rDNA and kDNA minicircles. We harnessed the CRISPR-Cas12a system as the detection method to avoid possible false positive results caused by non-specific amplification in the LAMP reaction and thereby improve the assay’s specificity. The assay process can be completed within 1.5 h, with results readable via either of the two optimized readouts: a fluorescence-based readout (suitable for laboratories with minimal molecular biology equipment such as a blue light transilluminator or a handheld fluorimeter) and a visual readout on lateral flow strips (to simplify the use of these assays). We report on the analytical validation and initial performance evaluation of our assays in clinical samples under controlled conditions in a research laboratory setting. With these achievements, compared to our previous proof-of-concept PCR-CRISPR assays [45], we aim to pave the way towards making CRISPR-based diagnostics accessible in a testing kit format for the diagnosis of leishmaniasis in rural endemic areas.

Here, in keeping with our *in silico* primer and crRNA design within conserved target genomic regions, the analytical validation demonstrated the ability of the 18S LAMP-CRISPR assay to achieve pan-*Leishmania* detection, whereas the kDNA LAMP-CRISPR assay reliably detected species belonging to the *Leishmania* (*Viannia*) subgenus. The kDNA assay also detected certain strains from the analyzed species of the *L*. (*Leishmania*) subgenus, specifically *L. mexicana* from Central America and *L. donovani* from the Old World. These results reflect both the similarity and variations in mitochondrial kDNA minicircle sequences between the two subgenera (see S1 Fig). Furthermore, our assays showed no cross-reactivity against other pathogens tested, which co-circulate in the same geographic regions and can thus coinfect the same human host, such as *T. cruzi* (a closely related trypanosomatid), *P. falciparum*, and *M. tuberculosis*, or cause cutaneous lesions similar to CL (*M. tuberculosis*) [81]. Our LAMP-CRISPR assays achieved high analytical sensitivity on serially diluted DNA from the *L. braziliensis* M2904 strain, with a detection level equivalent to 0.2 parasites per reaction. This detection sensitivity (below one parasite genome equivalent) compares well with that of the reference kDNA qPCR assay (5 × 10^-3^ genome equivalents per reaction; [64]), which is clinically relevant.

To explore the performance of our LAMP-CRISPR assays to detect *Leishmania* DNA in clinical samples, we evaluated a blinded panel of human skin lesion samples (n= 90) from patients enrolled in the Cusco region of Peru, where CL is caused by *L*. (*Viannia*) species. Both LAMP-CRISPR assays with a fluorescence readout attained good diagnostic performance, with sensitivity rates of 90.9% for kDNA and 72.7% for 18S rDNA, and a specificity of 100%, compared to the reference kDNA qPCR assay. Discrepancies between the two methods corresponded to samples with low parasite loads (S1 Dataset). Overall, LAMP-CRISPR assay results for the subgroup of samples tested with both fluorescence-based and lateral flow strip readouts were concordant. The 18S LAMP-CRISPR assay showed a somewhat reduced sensitivity when applied to clinical samples. This is likely related to the differences in target copy number: *Leishmania* parasites present an estimated 10 to 170 copies of the nuclear 18S rDNA region per genome [82, 83], while they harbor about 10,000 copies of extrachromosomal kDNA minicircles per cell [84].

Several studies have developed prototype LAMP assays for detecting *Leishmania* infections in diagnosing human VL, CL, and post-kala-azar dermal leishmaniasis (PKDL), as well as in domestic animal reservoirs (dogs) and sand flies. They target multicopy genomic regions, such as kDNA minicircles [85–87], the 18S rRNA gene [88–90], the ITS1 region in the rDNA array [91], and cysteine proteinase B (*cpb*) genes [92]. These assays proved to be highly sensitive for the detection of VL- and CL-causing *Leishmania* species (at the genus, species complex, or species level, depending on the target used), underscoring their potential value as diagnostic and surveillance tools in endemic settings. With the genus-specific targets, because of their high sequence conservation among the Trypanosomatidae, some nonspecific results were observed. For instance, the study of Adams *et al*. [88] reported on a LAMP assay based on 18S rDNA that was unable to discriminate between infections with *Leishmania*, *T. brucei*, and *T. cruzi*. Since the diseases caused by these related pathogens differ in clinical presentation, their differential diagnosis can be made on clinical symptoms and confirmed using a species-specific LAMP assay designed for this purpose [88]. In this study, when we tested our 18S LAMP primer set - crRNA combination, no cross-reactivity was observed with DNA from 3 *T. cruzi* strains belonging to different genetic lineages known as discrete typing units (DTUs), namely DTU TcI (Sylvio X10 strain), DTU TcIV (VT-1 strain), and DTU TcVI (Tulahuen LacZ clone C4 strain). Our results demonstrate the high analytical specificity of the LAMP-CRISPR assay towards *Leishmania* spp. We also noted that integrating LAMP amplification with downstream CRISPR-based detection improved discrimination between positive and negative amplification results, as shown in previous studies [37, 56]. This outcome is due to the LAMP-coupled CRISPR assays offering two layers of specificity: the first one enabled by using 4 to 6 LAMP primers that selectively target 6 to 8 distinct regions in the target DNA, and the second one due to the crRNA-guided Cas12a-mediated target sequence recognition [23]. An optimization to improve the LAMP assay specificity involves incorporating a probe-specific detection readout [18], as illustrated in the study by Ruang-Areerate *et al*. [93] aimed at detecting asymptomatic *Leishmania* infections in HIV-infected patients.

The LAMP technology is commercially available and delivered, such as the *Leishmania* detection kit (Loopamp™ *Leishmania* Detection Kit; Eiken Chemical Co., Ltd., Japan), which does not require cold-chain storage. This kit targets kDNA minicircles and 18S rDNA in the same reaction to enable pan-*Leishmania* detection based on the LAMP assay developed by Adams *et al*. [94]. The diagnostic performance evaluation of this kit in different endemic areas around the world has demonstrated its high sensitivity and specificity for the diagnosis of VL [22, 76, 94–96], CL [22, 94, 97, 98], and PKDL [99], underscoring its potential for implementation in primary healthcare facilities. Undoubtedly, the DNA extraction method can impact LAMP assay performance when testing clinical samples. As shown in the study by Ghosh *et al*. [99], a lower efficiency of LAMP to diagnose PKDL was observed when the LAMP reaction was performed using crude DNA obtained by a simple in-house boil and spin protocol compared to DNA extracted by a commercial silica column-based (Qiagen) kit (67.2% vs. 89.7% sensitivity, respectively). Further, a recent study found a compromised sensitivity (48.4%) of the *Leishmania* LAMP kit for the diagnosis of CL in Ethiopia, possibly due to a primer mismatch with the *L. aethiopica* 18S rDNA target [100]. This finding points out the need to extensively validate existing LAMP assays in diverse clinical contexts and epidemiological scenarios in different geographical areas to confirm their effectiveness in detecting local parasite variants that are circulating. The same applies to our new LAMP-CRISPR assays.

Combining highly sensitive isothermal amplification with the notable highly specific CRISPR-Cas12a system is an approach that has been applied for detecting pathogenic trypanosomatids. For example, Wiggins *et al*. [46] developed proof-of-concept RPA-coupled CRISPR-Cas12a assays that target kDNA maxicircles for *Leishmania* spp. detection. They have recently adapted their assays into a multichambered device to obtain a rapid nucleic acid diagnostic tool that does not require electricity or instrumentation, a promising development for diagnostic applications in low-resource settings [101]. In another study, Yang *et al*. [47] developed a one-tube RPA-CRISPR/Cas12a assay that targets the *Leishmania Kmp11* gene. The assay process can be completed within 50 min, with the fluorescent signal of reaction tubes observed visually under blue light illumination. Their assay has been evaluated on simulated clinical samples, yielding a sensitivity of 97.9% and a specificity of 100%. Ortiz-Rodríguez *et al*. [50] developed a platform for *T. cruzi* detection, which was validated on samples from reservoirs and insect vectors. It employs PCR or RPA followed by CRISPR-Cas12a-based detection. Of 3 nuclear target genes evaluated, namely cytochrome B (*Cytb*), the 18S rRNA gene, and histone H2A, only the former (*Cytb*) resulted in a workable CRISPR-based assay since no mutations were found in the chosen target site during testing of *T. cruzi* DNA extracted from the intestine of triatomine vectors. Furthermore, the authors designed a portable prototype device (called TropD-Detector) for fluorescence results discrimination, suitable for low-resource settings. Guarnizo *et al*. [56] designed a LAMP-CRISPR/Cas12a assay that targets a highly conserved region in the nuclear *HSP70* gene to specifically detect *T. cruzi* in the context of acute Chagas disease diagnosis in endemic countries. They evaluated the assay in clinical samples from 100 infants born to Chagas-positive mothers in Bolivia, achieving a sensitivity of 77.27% and a specificity of 100% compared to a TaqMan-based qPCR assay that targets satellite DNA.

Our study shows that LAMP-coupled CRISPR-Cas12a assays are an effective alternative method for detecting *Leishmania* infections in different non-invasive and invasive specimens (S1 Dataset) from human skin lesions. While kDNA LAMP primers previously reported were for a *L. donovani* complex-specific assay [94] or for a *L. amazonensis*-specific assay [87], our kDNA LAMP-CRISPR assay (specifically the crRNA design) exploited a kDNA minicircle region conserved in the New World species of the *Leishmania* (*Viannia*) subgenus ([45] and S1 Fig). Our pan-*Leishmania* 18S LAMP-CRISPR assay is expected to be capable of detecting various parasite species from both the New and Old World (S2 Fig). Our findings encourage further evaluation and validation of both assays in different geographical areas. Besides, our assays have the potential to be adapted in future work to address the ASSURED/REASSURED criteria for a diagnostic test suitable for adoption across different healthcare levels, which require assays to be affordable, sensitive, specific, user-friendly, robust and rapid, as well as equipment-free (or operating with minimal equipment), deliverable to those who need them, with real-time connectivity, and workable on easily collected specimens [102, 103]. Sample preparation is one of the challenges in molecular assay implementation in resource-limited settings [104]. Further research is needed to optimize rapid DNA extraction protocols, such as boil-spin [99] and SpeedXtract (a magnetic bead-based method) [105], combined with a streamlined CRISPR assay workflow to move forward to a near-PoC nucleic acid-based diagnostic assay for CL and other forms of leishmaniasis. Both LAMP and CRISPR reactions are compatible with freeze-drying (lyophilization), which improves reagent stability for easy transport and storage at room temperature and may reduce contamination risks [106]. This improvement can also facilitate the preparation of the assays in a pre-designed kit format, enabling a portable assay setup for resource-limited settings. Therefore, the impact of freeze-drying reagents on the performance of our assays needs to be evaluated in future work. Incorporating these refinements into our assay procedure, along with user-friendly devices, such as the LFA strips or a handheld fluorimeter, would allow its adoption in minimally-equipped laboratories in endemic regions, and may be incorporated in a mobile suitcase laboratory. The latter approach has been explored with a RPA assay in field studies conducted in Asia, showing promising performance for VL, PKDL, and CL diagnosis in endemic settings [105, 107, 108]. After further optimization and validation, we envisage that our LAMP-CRISPR assays may become valuable tools for providing rapid testing results directly in the affected regions, thereby mitigating delays associated with testing in centralized laboratories. CRISPR-based assays could also have a considerable role in epidemiological studies, in providing real-time surveillance data. Further, an approach for future work that offers the potential to be impactful in resource-limited endemic settings entails the integration of our assays with recent developments in lab-on-chip systems, such as a paper-based LAMP-CRISPR integrated diagnostic platform (PLACID) [37]. This paper-based device uses freeze-dried LAMP and CRISPR reagents, where nucleic acid amplification and CRISPR reactions are performed successively without fluid transfer using paper microfluidics. A smartphone-based protocol allows the fluorescent detection of nucleic acid targets. This platform showed robust performance for bacterial and viral pathogen target detection, with no false positives observed [37]. These are well-known refinements to consider in future assay development efforts. Indeed, we noticed that our current two-step LAMP-CRISPR open assay format intrinsically carries the risk of cross-contamination during sample transfer from tube-based (LAMP) to plate-based (CRISPR) reactions, as reported in other studies [106].

In conclusion, we developed novel LAMP-CRISPR assays for the molecular detection of *Leishmania*, targeting two multicopy genomic regions: 18S rDNA for pan-*Leishmania* detection and a kDNA minicircle region conserved among New World *L*. (*Viannia*) species. By integrating LAMP with CRISPR-Cas12a technology, the assays enable accurate detection, with results readout via fluorescence or lateral flow strips, the latter aimed at facilitating a simpler workflow. Analytical validation and initial performance testing on clinical samples demonstrated high sensitivity and specificity, underscoring the assays’ potential as effective alternatives for detecting *Leishmania* infections. With further optimization and validation, these prototype assays hold promise as next-generation molecular tools with great potential to improve the diagnosis and surveillance of leishmaniasis in endemic regions, thereby supporting One Health strategies for disease control.

## Data Availability

All relevant data are within the manuscript and its Supporting Information files.

## Supporting information captions

**S1 Fig. Multiple sequence alignment of representative kDNA minicircle sequences from members of the *L*. (*Viannia*) and *L*. (*Leishmania*) subgenera, with LAMP primer binding regions and crRNA target site highlighted.** A total of 24 *L*. (*Viannia*) and 35 *L*. (*Leishmania*) sequences were aligned using Clustal Omega (alignments were performed separately for each subgenus) and visualized in Jalview v2.11.4.1. The locations of primer binding regions as well as of the crRNA target site and PAM sequence in the target DNA are highlighted with colors to indicate sequence conservation within the *L*. (*Viannia*) subgenus and sequence divergence within the *L*. (*Leishmania*) subgenus. Nucleotide variants are represented by spaces in the alignment. The kDNA minicircle sequence from the *L.* (*V.*) *braziliensis* M2904 strain (GenBank accession no. KY698803.1) exhibited 100% identity across all primer and crRNA target regions. The sequences shown are from selected representative strains/isolates of a given *Leishmania* species; additional species and sequences can be examined in the full alignments included in the S1 File. Figure created with BioRender.com.

**S2 Fig. Multiple sequence alignment of representative 18S rDNA sequences from members of the *Leishmania* and *Trypanosoma* genera, with LAMP primer binding regions and crRNA target site highlighted.** A total of 31 sequences (16 from *Leishmania* spp. and 15 from *Trypanosoma* spp.) were aligned using Clustal Omega and visualized in Jalview v2.11.4.1. Primer binding regions as well as the crRNA target site and PAM sequence in the target DNA are highlighted with colors to indicate sequence conservation within the *Leishmania* genus and sequence variations within the *Trypanosoma* genus. Nucleotide variants are represented by spaces in the alignment. The sequences shown are from selected representative strains/isolates of a given species; additional species and sequences can be examined in the full alignments included in the S2 File. Figure created with BioRender.com.

**S3 Fig. Optimization of the incubation time of the LAMP reaction**. To determine the minimum necessary incubation time of the LAMP reaction for consistent DNA amplification, tests were performed at different reaction times for *Leishmania* kDNA (10, 20, 30, and 40 min) and 18S rDNA (10, 30, 40, 50, and 60 min) targets. The number and text enclosed in a rectangle indicates the LAMP reaction time. The amplification efficiency was assessed using varying amounts of template DNA (*L. braziliensis* M2904 gDNA), corresponding to approximately 2 × 10^2^, 2 × 10^-1^, and 2 × 10^-3^ genome equivalents. A negative control (NTC) reaction was tested in parallel. Raw fluorescence signals from Cas12a reactions are shown for kDNA (A-D) and 18S rDNA (G-K) for one representative technical replicate. Normalized data (fluorescence ratio at *t* = 25 min) from two technical replicates are shown in panels E (kDNA) and L (18S rDNA). A fluorescence ratio ≥ 2 is considered detected. Fluorescence measurements in this figure were taken on the Varioskan LUX plate reader. LAMP reaction products (5 µL each) of kDNA (F) and 18S rDNA (M) were analyzed by 2% agarose gel electrophoresis ran at 100 V for 1 h using SYBR Gold staining. L, 100 bp DNA ladder (ABclonal). Since space was tight on the agarose gel images, we had to abbreviate the labels for each lane. For instance, “10^2^” actually represents “2 × 10^2^” as indicated in other panels of this figure.

**S4 Fig. Optimization of CRISPR-based detection assays using a lateral flow readout**. (A) Design of the Milenia GenLine HybriDetect test strips and readout interpretation. (Left) In the absence of the specific genetic target, the intact biotin-FAM labeled reporter molecule flows to the control line (C-line). This is interpreted as a negative test result. (Right) Upon recognition of the genetic target, the CRISPR RNP complex cleaves the reporter molecule, which flows to the test line (T-line). The signal intensity of the C-line is weakened. If cleavage of the reporter molecule is partial, both the C-line and T-line may appear with comparable intensity. Both scenarios are interpreted as a positive test result. Figure adapted from [109] and created with BioRender.com. (B-F) Different parameters influencing the LFA readout performance were evaluated to achieve consistent results: reporter concentration (B), assay buffer (C), incubation temperature (D) and reaction time (E) of the Cas12a assay, and the concentration of PEG-8000 (F). Results shown here correspond to the kDNA LAMP-CRISPR assay.

**S5 Fig. Workflow from sample preparation to LAMP-CRISPR assays for analytical specificity testing**. Figure created with BioRender.com.

**S6 Fig. Results of LAMP-CRISPR detection with fluorescence readout in a group of clinical samples**. Raw fluorescence curves generated by Cas12a detection of LAMP amplicons from *Leishmania* kDNA (A) and 18S rDNA (B) targets in a representative group of clinical samples harboring varying parasite load levels (see S1 Dataset) and ran in the same assay plate per genomic target. Out of the 8 tested samples shown, 6 showed robust fluorescence curves indicating the presence of *Leishmania* kDNA molecules (panel A; samples coded S16, S23, S31, S52, S72, and S75) whereas the 18S rDNA target was detected in 4 of them (panel B; samples coded S16, S23, S52, and S75). Fluorescence measurements in this figure were taken on the Synergy H1 plate reader. The respective LAMP amplicons (5 µL) targeting kDNA (C) or 18S rDNA (D) were analyzed by 2% agarose gel electrophoresis (100 V, 60 min) using SYBR Gold staining. Controls included a positive control (PC, 5 × 10^4^ genome equivalents from *L. braziliensis* M2904 gDNA), a human negative control (HC, human PBMC gDNA), and a no-template control (NTC) of the LAMP reaction.

**S7 Fig. Results of LAMP-CRISPR detection with LFA readout in a group of clinical samples**. Clinical samples chosen for this pilot testing (n = 8) were categorized according to the parasite load levels and tested with kDNA and 18S LAMP-CRISPR assays. Lateral flow testing was performed on two assay repeats, R1 and R2, per sample. Visualization on the lateral flow strips (Milenia GenLine HybriDetect strips) was achieved through the cleavage of the biotin-FAM reporter molecule. Following LFA analysis, the strips were placed into the cassette housing for image acquisition. C, control line; T, test line. Controls included DNA samples from two non-leishmaniasis patients (i.e., kDNA qPCR-negative; samples 09 and C7), a positive control (PC, 2 × 10^2^ genome equivalents from *L. braziliensis* M2904 gDNA), a human negative control (HC, 20 ng of human PBMC gDNA), and a no-template control (NTC) of the LAMP reaction.

**S1 Table. Primer and crRNA template sequences used in this study.**

S1 File. *In silico* alignment of *Leishmania* kDNA minicircle sequences from *Viannia* and *Leishmania* subgenera, and location of primer and crRNA binding sites. This file contains sequence alignments and visualizations used to assess the *in silico* specificity of the designed LAMP primers and Cas12a crRNA within conserved regions of *Leishmania* kDNA minicircles. A total of 1,060 aligned sequences from strains/isolates belonging to the *L*. (*Viannia*) subgenus and 965 aligned sequences from strains/isolates of the *L*. (*Leishmania*) subgenus were included. Primer and crRNA binding regions were manually annotated using Jalview v2.11.4.1 to enhance visualization and facilitate identification of conserved target sites. Additionally, a subset of representative sequences, 24 *L*. (*Viannia*) and 35 *L*. (*Leishmania*), is provided in separate alignment project files to illustrate kDNA minicircle sequence conservation and divergence across the two subgenera (see also S1 Fig).

**S2 File. *In silico* alignment of 18S rDNA sequences from *Leishmania* and *Trypanosoma* genera, and location of primer and crRNA binding sites.** This file contains the alignments and visualizations used to evaluate the *in silico* specificity of the designed LAMP primers and Cas12a crRNA targeting the 18S rDNA. It includes 37 aligned sequences of *Leishmania* spp. and 272 of *Trypanosoma* spp. Primer and crRNA binding regions were manually annotated using Jalview v2.11.4.1 to enhance visualization and identification. Additionally, an alignment of a subset of 31 representative sequences (16 *Leishmania*, 15 *Trypanosoma*) is provided for visualization of 18S rDNA sequence conservation and divergence across the two genera (see also S2 Fig).

**S1 Dataset. Excel file containing the data on tested clinical samples generated during the current study**. This file contains the LAMP-CRISPR and qPCR data on tested clinical samples, as well as the determination of threshold cutoff values for fluorescence-based CRISPR-Cas12a assay result interpretation.

## Author Contributions

Conceptualization: Vanessa Adaui.

Formal analysis: Eva Dueñas, Percy Huaihua, Vanessa Adaui. Funding acquisition: Vanessa Adaui.

Investigation: Eva Dueñas, Ingrid Tirado, Percy Huaihua, Ariana Parra del Riego, Luis Cabrera-Sosa, Jose A. Nakamoto, Carlos M. Restrepo.

Project administration: Vanessa Adaui.

Resources: Luis Cabrera-Sosa, Jose A. Nakamoto, María Cruz, Carlos M. Restrepo, Jorge Arévalo, Vanessa Adaui.

Supervision: Vanessa Adaui, Jorge Arévalo. Visualization: Eva Dueñas, Ingrid Tirado.

Writing – original draft: Eva Dueñas, Ariana Parra del Riego, Vanessa Adaui.

Writing – review & editing: Eva Dueñas, Ingrid Tirado, Percy Huaihua, Ariana Parra del Riego, Luis Cabrera-Sosa, Jose A. Nakamoto, María Cruz, Carlos M. Restrepo, Jorge Arévalo, Vanessa Adaui.

## Acknowledgments

We thank Pohl Milón (Universidad Peruana de Ciencias Aplicadas) for inspiring discussions; Yomara Romero (Universidad Peruana de Ciencias Aplicadas) for assistance with sequence alignments and advice on their presentation; Edith Málaga and Manuela Verástegui (Universidad Peruana Cayetano Heredia) for providing genomic DNA from the *Trypanosoma cruzi* strains tested here; and the Office of Research at the Universidad Peruana de Ciencias Aplicadas for the support provided to carry out this research work.

