## Supplementary figures and images for "LAMP-coupled CRISPR-Cas12a assays for upgrading molecular detection of *Leishmania* infections"

### Supplemental Figure 1

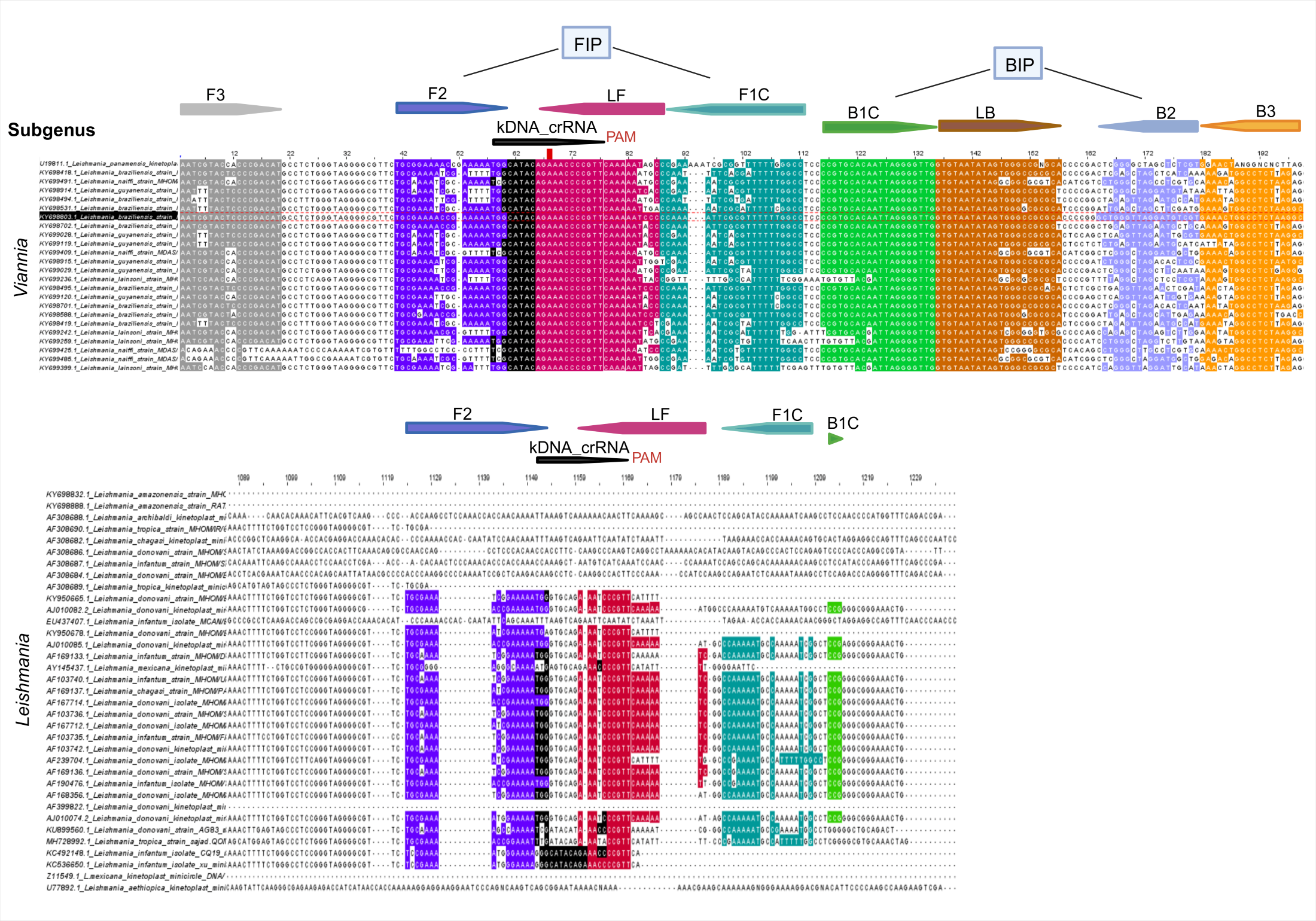

### Supplemental Figure 2

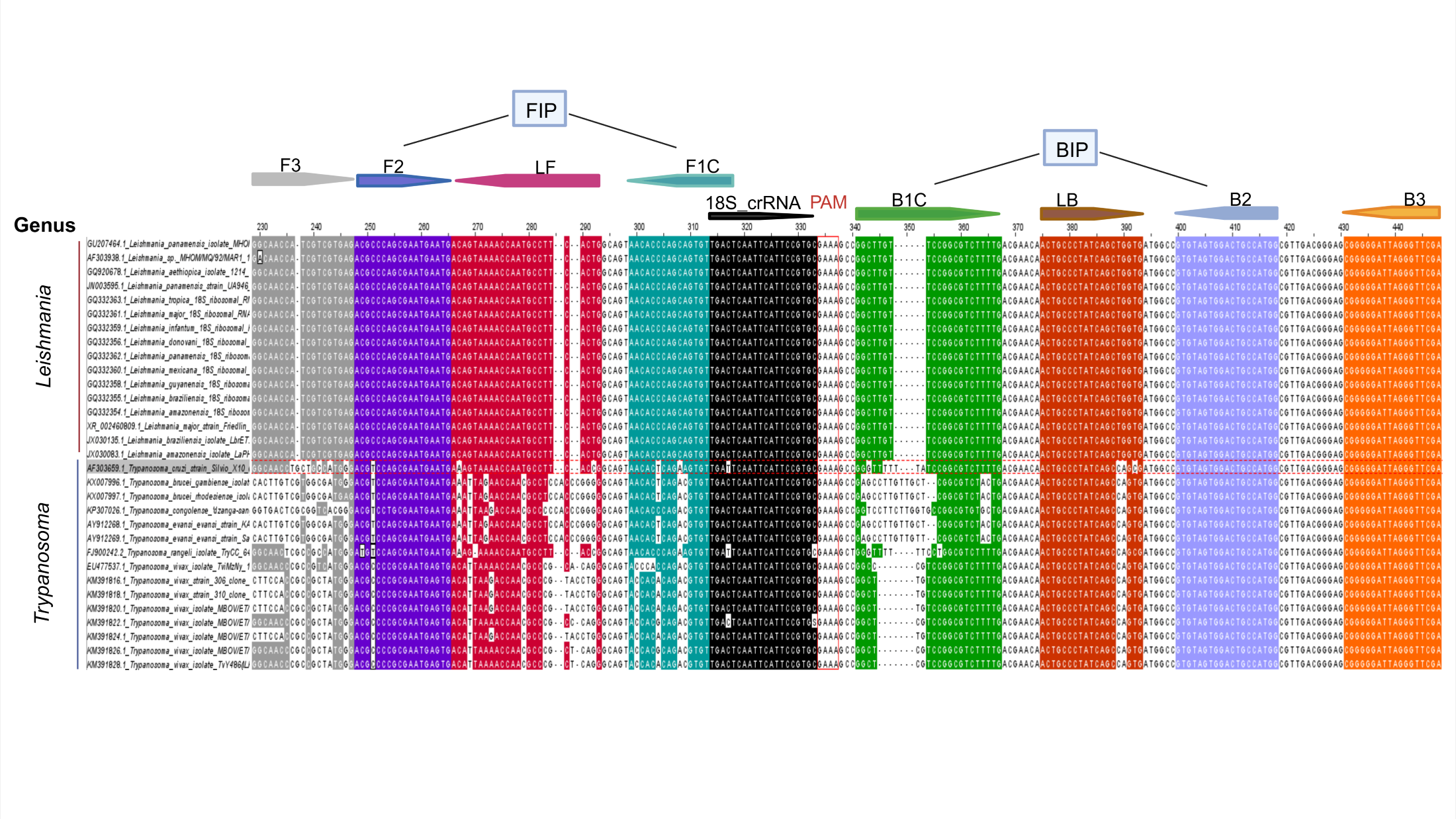

### Supplemental Figure 3

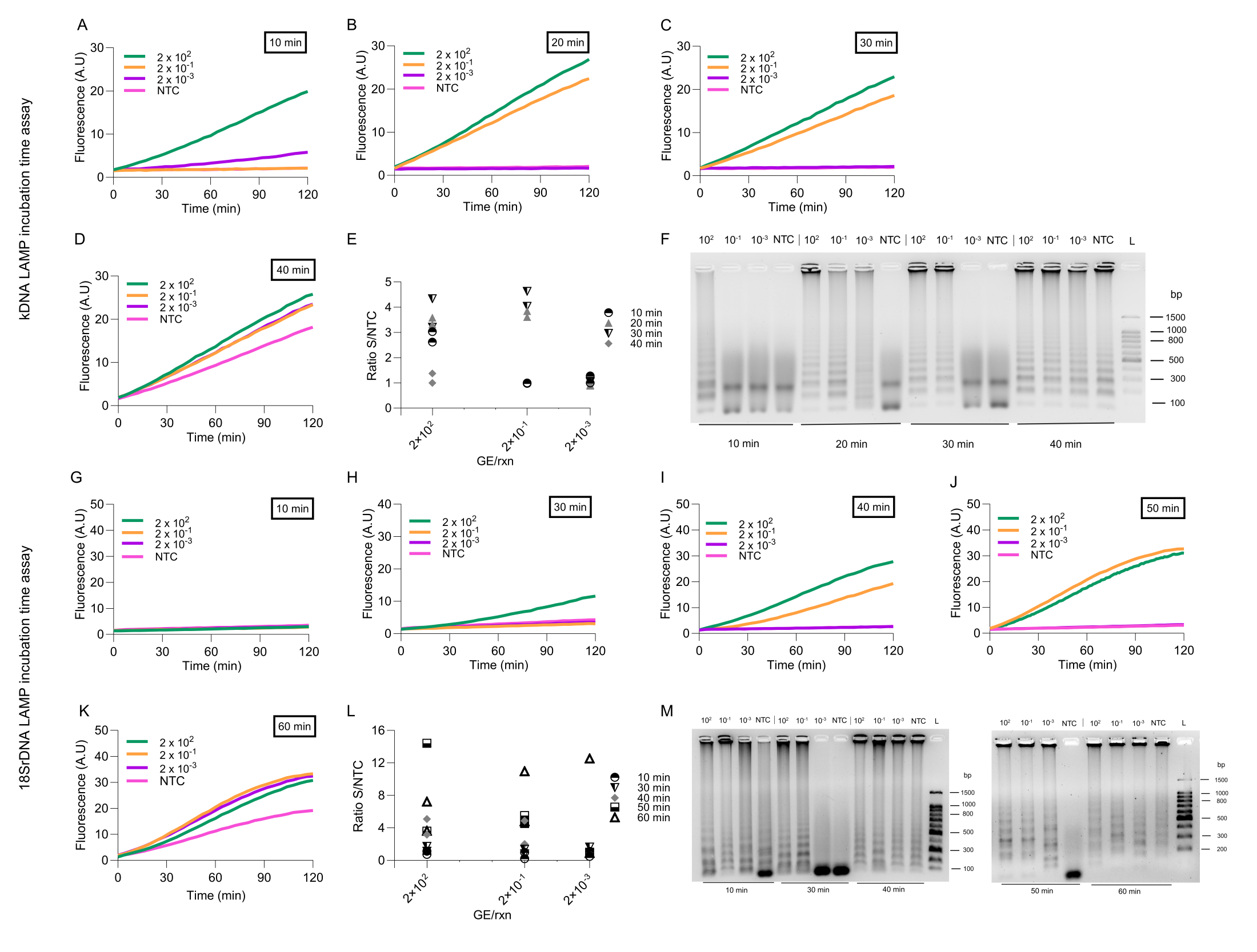

### Supplemental Figure 4

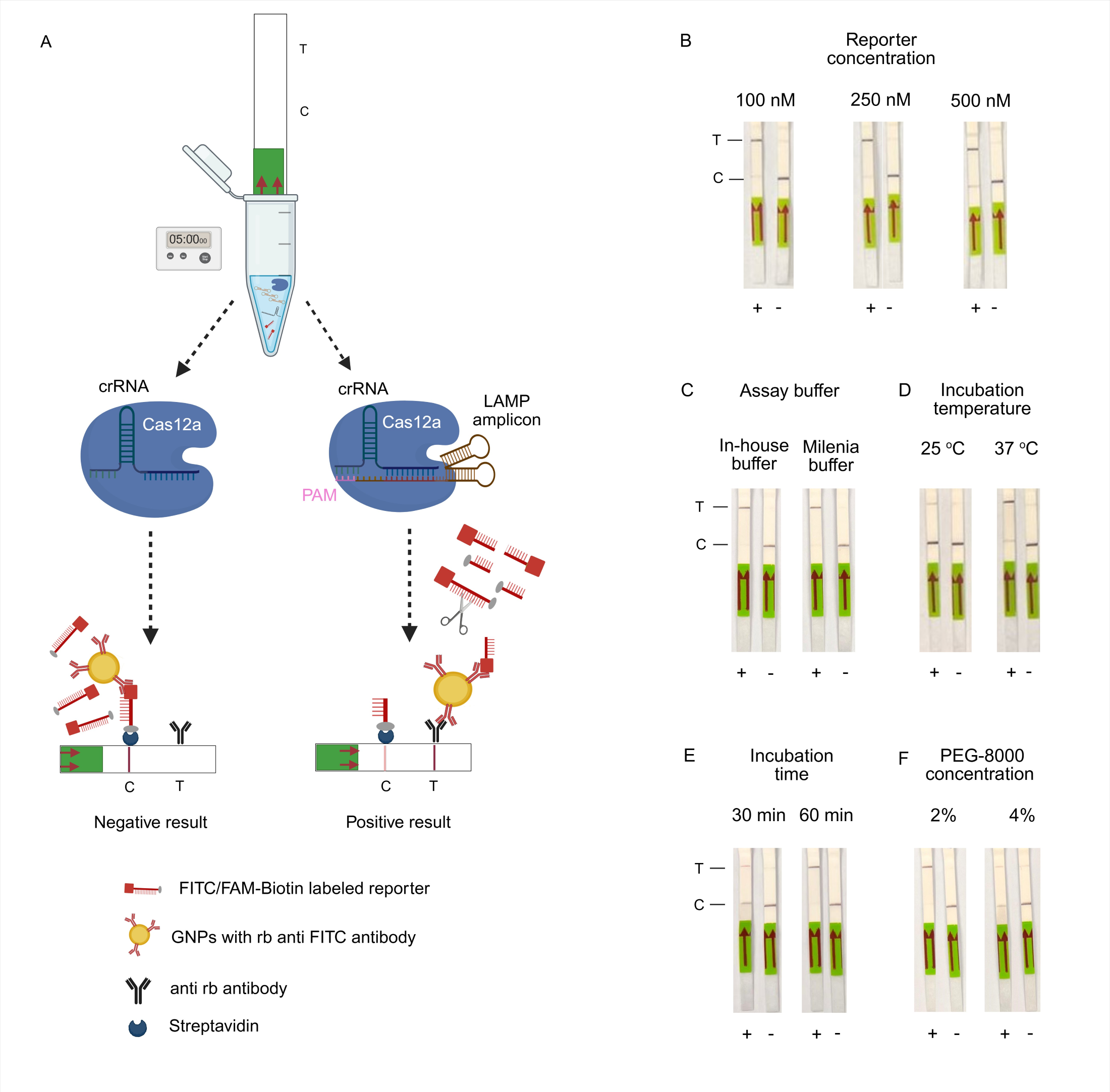

### Supplemental Figure 5

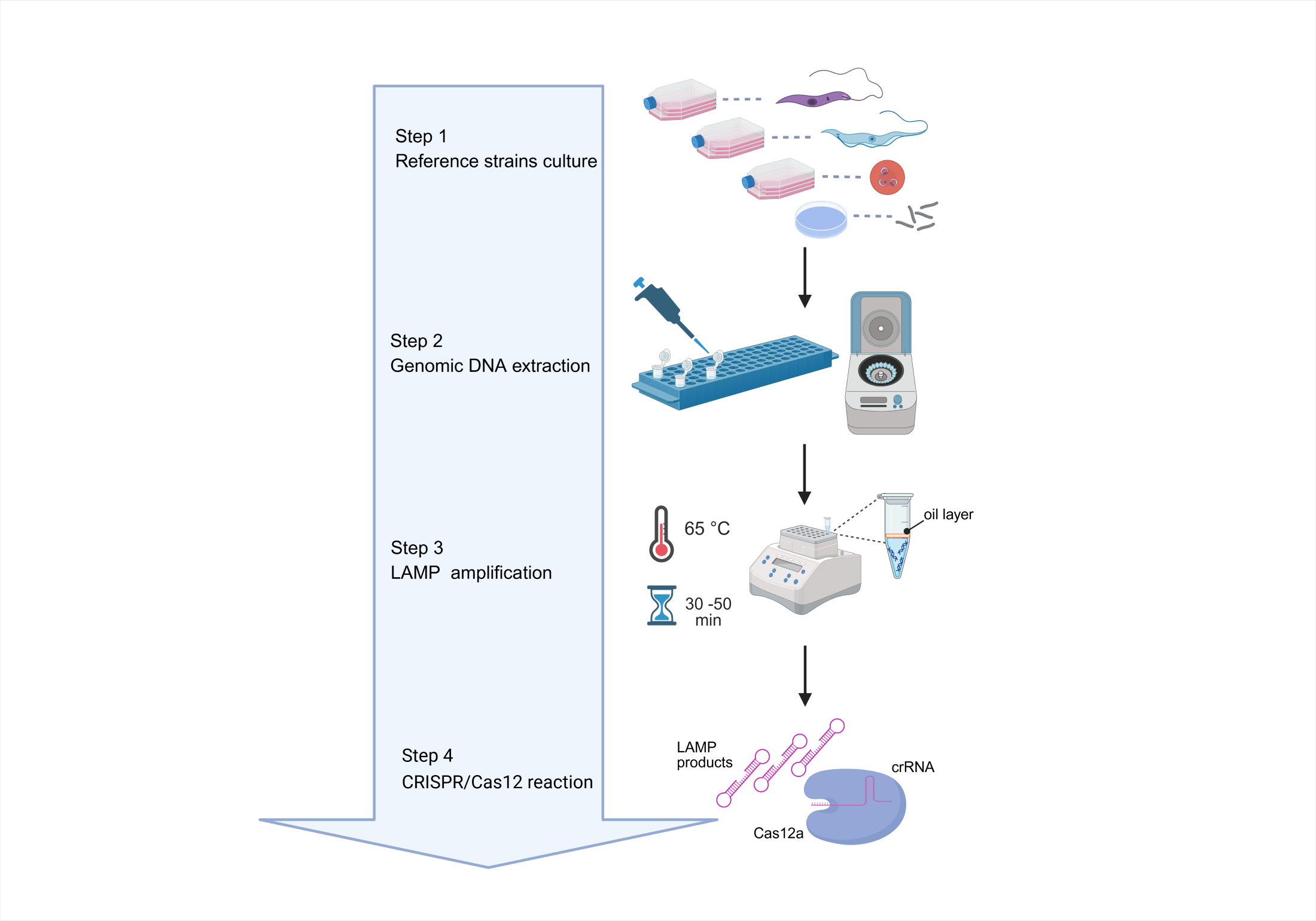

### Supplemental Figure 6

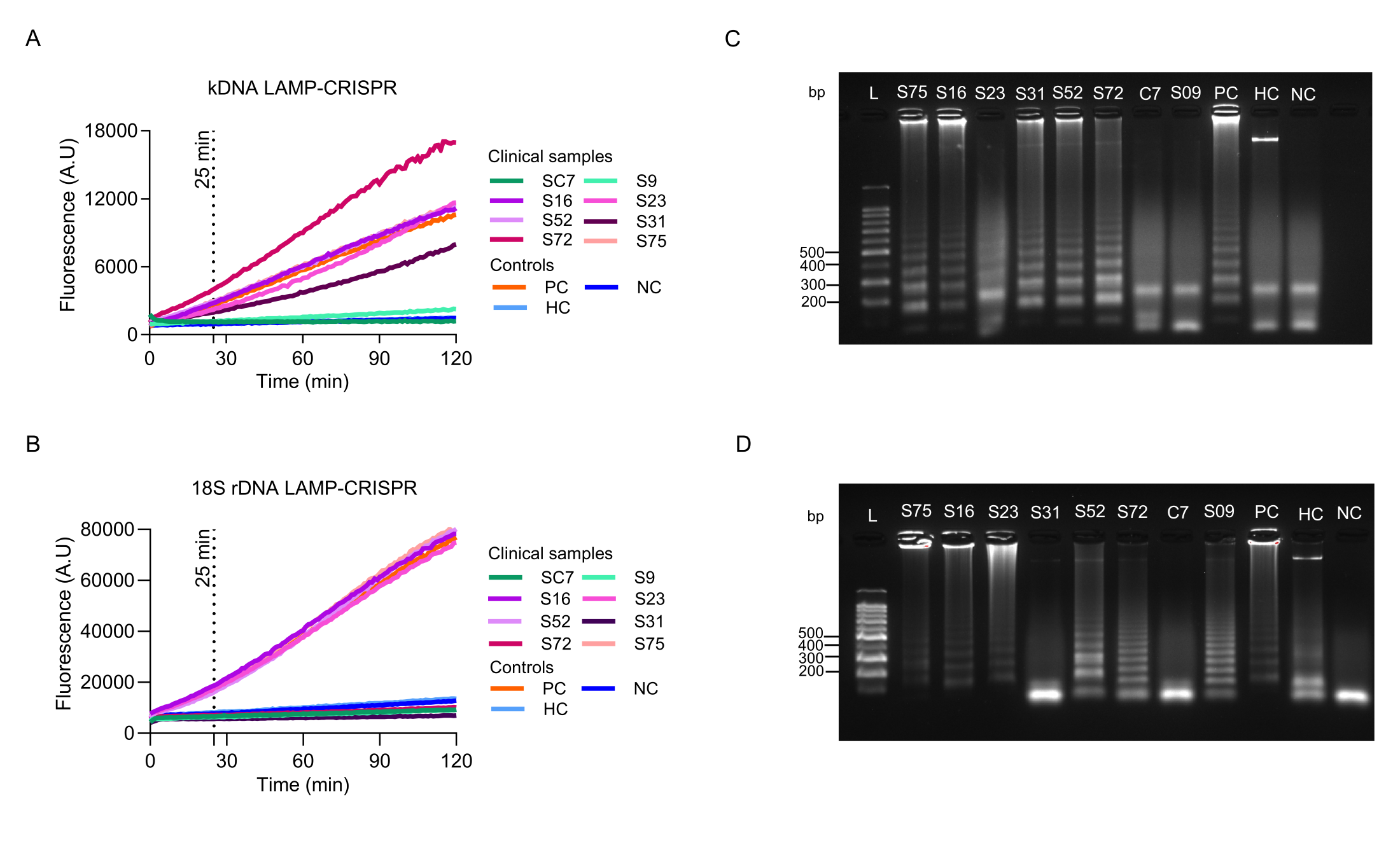

### Supplemental Figure 7

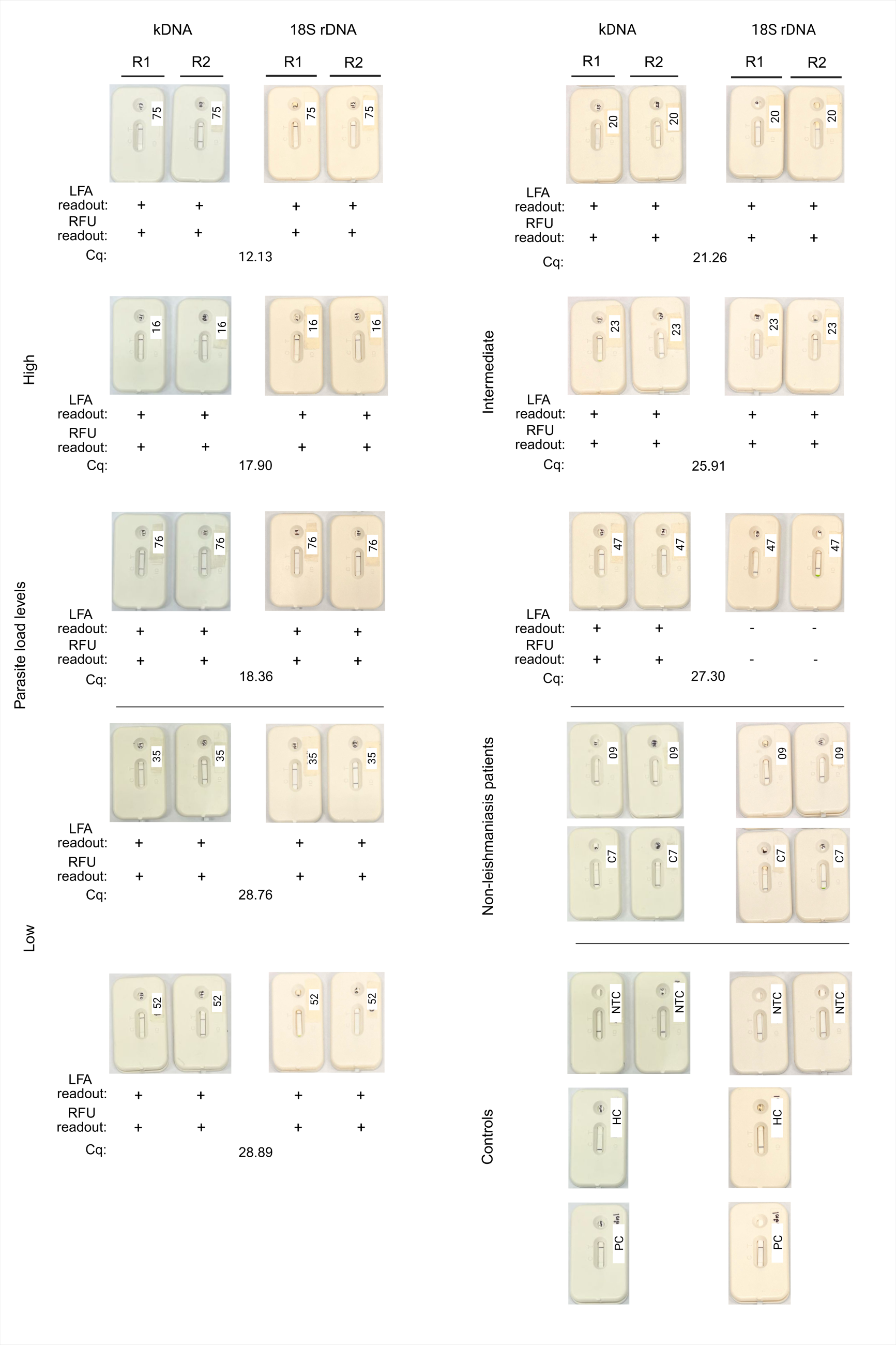
