## Supplemental Table 1 for "LAMP-coupled CRISPR-Cas12a assays for upgrading molecular detection of *Leishmania* infections"

S1 Table. Primer and crRNA template sequences used in this study.

| ID | Sequence 5' - 3' | Description | Application ¥ | Reference |
|---|---|---|---|---|
| 18S_crRNA | TAATACGACTCACTATAGGtaatttctactaagtgtagat GCACGGAATGAATTGAGTCA | 18S rDNA dsDNA template (*) | 18S crRNA synthesis | Dueñas et al., 2022 |
| kDNA_crRNA | TAATACGACTCACTATAGGtaatttctactaagtgtagat AACGGGGTTTCTGTATGCCA | kDNA minicircle dsDNA template (*) | kDNA minicircle crRNA synthesis | Dueñas et al., 2022 |
| 18S_FIP | TGAGTCAACACTGCTGGGTGTTACGCCCAGCGAATGAATG | 18S forward internal primer | 18S target pre-amplification | This study |
| 18S_BIP | GGCTTGTTCCGGCGTCTTTTGCCATGGCAGTCCACTACAC | 18S backward internal primer | 18S target pre-amplification | This study |
| 18S_F3 | GGCAACCATCGTCGTGAG | 18S forward external primer | 18S target pre-amplification | This study |
| 18S_B3 | TCGAACCCTAATCCCCCG | 18S backward external primer | 18S target pre-amplification | This study |
| 18S_LF | CAGTGAAGGCATTGGTTTTACTGT | 18S forward loop primer | 18S target pre-amplification | This study |
| 18S_LB | ACTGCCCTATCAGCTGGTG | 18S backward loop primer | 18S target pre-amplification | This study |
| kDNA_FIP | GGCCAAAAAACGCGAATTTTGGTGCGAAAACCGAAAAATGG | kDNA forward internal primer | kDNA target pre-amplification | This study |
| kDNA_BIP | CCGTGCACAATTAGGGGTTGACGACATCCTAACCCAGC | kDNA backward internal primer | kDNA target pre-amplification | This study |
| kDNA_F3 | AATCGTACTCCCCGACAT | kDNA forward external primer | kDNA target pre-amplification | This study |
| kDNA_B3 | GCCTTAGAGGCCAGTTTC | kDNA backward external primer | kDNA target pre-amplification | This study |
| kDNA_LF | GGGATTTTTGAACGGGGTTTCT | kDNA forward loop primer | kDNA target pre-amplification | This study |
| kDNA_LB | GTGTAATATAGTGGGCCGCGCA | kDNA backward loop primer | kDNA target pre-amplification | This study |
| MP1L | TACTCCCCGACATGCCTCTG | kDNA forward qPCR primer | kDNA qPCR | López et al., 1993; Jara et al., 2013 |
| MP3H | GAACGGGGTTTCTGTATGC | kDNA reverse qPCR primer | kDNA qPCR | López et al., 1993; Jara et al., 2013 |
| PHP10-F | CATGGGAAGCAAGGGAACTAATG | ERV-3 forward qPCR primer | Human ERV-3 qPCR, for normalization of the parasite load | Yuan et al., 2001 |
| PHP10-R | CCCAGCGAGCAATACAGAATTT | ERV-3 reverse qPCR primer | Human ERV-3 qPCR, for normalization of the parasite load | Yuan et al., 2001 |

(*) The dsDNA templates for crRNA generation through *in vitro* transcription were designed with a T7 promoter (underlined sequence), followed by the scaffold sequence (lowercase letter), and the specific recognition sequence (uppercase letter).

Abbreviations: rDNA, ribosomal DNA; dsDNA, double-stranded DNA; kDNA, kinetoplast DNA; ERV-3, endogenous retrovirus 3; crRNA, CRISPR RNA.
¥ Target pre-amplification refers to the LAMP amplification step prior to Cas12a-based detection.
